# Whole-of-School Physical Activity Implementation in the Context of the Dubai Fitness Challenge

**DOI:** 10.1101/2023.08.17.23294245

**Authors:** Chris McMahon, Collin A. Webster, R. Glenn Weaver, Christophe El Haber, Gönül Tekkurşun Demir, Zainab Mohamed Ismail, Syeda Zoha Fatima Naqvi, Mehnaz Ghani, Şevval Kepenek, Manel Kherraf, Thrisha Krishnakumar, Pranati Prakash, Yeowon Seo

## Abstract

**Introduction:** Physical activity (PA) promotion among school-aged youth is a global health priority. Recommendations for such promotion include implementing whole-of-school approaches that maximize resources across the school environment. This study examined schools’ participation in an annual, government-led, and emirate-wide initiative in Dubai, called the Dubai Fitness Challenge, in which the goal is to accrue 30 minutes of PA every day for 30 days (as such, the initiative is colloquially referred to as “Dubai 30×30”).

**Methods:** A mixed-methods design was employed for this study. Three schools were recruited using convenience sampling. Participants were 18 physical education teachers, 20 classroom teachers, 2 principals and 45 students. Data sources included surveys, focus groups, and interviews. Data were analyzed using descriptive statistics, multinomial logistic regression, and open and axial coding to develop themes.

**Results:** School staff reported that most Dubai 30×30 activities were provided in physical education, at break times during school, and before and after school. Students reported that they mainly participated in Dubai 30×30 activities during physical education and occasionally participated in activities after school and on weekends. During school, students were more likely to reach higher PA intensity levels when they were in contexts other than the regular classroom setting. Among school staff, physical education teachers were most involved and classroom teachers were least involved in promoting Dubai 30×30. Parent engagement was high. Staff perceived that Dubai 30×30 brought the community together, but physical education teachers also indicated there was a lack of implementation guidance and they felt burdened. Participants believed Dubai 30×30 increased PA participation and helped to promote their schools.

**Discussion:** This study provides an initial glimpse into schools’ participation in Dubai 30×30 and suggests that a whole-of-school PA lens is useful in gleaning information that could help to increase and optimize PA opportunities for students.

## Introduction

Physical activity (PA) benefits children and adolescents by supporting their physical, mental, social, and emotional development (Chaput et al., 2020). The World Health Organization (WHO) recommends that school-age youth engage in at least 60 minutes of moderate-to-vigorous intensity PA each day (WHO, 2020). This PA should be mostly aerobic and should include activities that strengthen muscles and bones at least three days per week (WHO 2020). Unfortunately, 81% of adolescents do not meet PA guidelines (Guthold et al., 2020). As physical inactivity can track from adolescence into young adulthood (Hayes et al., 2019; Telama, 2009), early intervention during the school years should be a priority for addressing insufficient PA levels in youth and promoting active living.

Given their extensive reach and existing resources, schools provide an attractive setting for supporting children and adolescents’ PA needs. Recommendations stress the importance of taking a “whole-of-school approach” to PA promotion (Global Advocacy for Physical Activity, 2012; Institute of Medicine [IOM], 2013; Milton et al., 2021). Such an approach may look different from one school to the next but the general idea is to maximize PA opportunities and participation by using as many resources as possible before, during and after school. For example, the comprehensive school physical activity program (CSPAP) framework, which is the national framework for school-based physical education and PA in the United States, identifies five components that can be used synergistically to help ensure all school-aged youth meet the PA guidelines and develop the competence and confidence to lead physically active lives (Centers for Disease Control and Prevention, 2019). These components include (a) quality physical education, (b) PA during school, (c) PA before and after school, (d) staff involvement and (e) family and community engagement. A quality physical education program should serve a foundational role in a CSPAP by providing students with the knowledge and skills to pursue a physically active lifestyle. Other PA opportunities during school (e.g., recess and other break times, PA integrated with classroom time), as well as PA opportunities before and after school (e.g., clubs, active transportation), can allow students to apply and extend what they learn in physical education. The staff involvement and family and community engagement components focus on recruiting the support of as many other people as possible (e.g., all teachers, school administrators, parents, community partners) to assist with PA promotion.

Evidence suggests that a whole-of-school approach to PA promotion leads to positive outcomes for school-age youth (Belton et al., 2020; Colabianchi et al., 2015; Katewongsa et al., 2022; Pulling Kuhn et al., 2021). For example, Pulling Kuhn et al. (2021) conducted a systematic review of multicomponent school-based PA interventions and found that reported outcomes included increased time spent in PA, increased physical fitness, improved motor skills, improved weight status, improved blood pressure, improved on-task behavior and improved performance in reading and math. However, surveys conducted in the U.S. indicate whole-of-school PA programs exist in only 16% of elementary schools and 1-7% of secondary schools (American Alliance for Health, Physical Education, Recreation and Dance [AAHPERD], 2011; Brener et al., 2017; Colabianchi et al., 2015). In Ireland, a national initiative with primary (elementary) schools called Active School Flag has had more success with 85% of schools engaging at some level with the initiative and 46% of schools earning at least one flag (i.e., meeting criteria to be named an “active school”).

The varying uptake of whole-of-school PA underscores the importance of examining schools’ implementation of current initiatives. Carson et al. (2014) proposed a conceptual framework for research and practice related to whole-of-school PA, which is based on ecological systems theory (Bronfenbrenner, 1992; McLeroy et al., 1988). The framework is organized into multiple levels of influence on the implementation of a whole-of-school PA initiative and, ultimately, on youth PA. Specific to implementation, the most proximal level of influence is the Meso level. This level consists of implementation facilitators, including the knowledge, skills, and dispositions of the implementers (e.g., school personnel), as well as resources (e.g., school facilities, equipment, time, and space allocation) and safety (physical, social, and emotional) regarding PA opportunities. Above the Meso level is the Exo level, which comprises the leaders of whole-of-school PA initiatives: a champion (someone who spearheads the initiative), the school administrative team (i.e., their support for the initiative) and a school committee dedicated to the initiative. Finally, the Macro level incorporates cultural elements of the school in relation to PA promotion. These elements include policy (e.g., PA-related legislation, monitoring and evaluation of the initiative) and normative behaviors and beliefs (e.g., shared expectations and values of members of the school community).

Research on whole-of-school PA implementation has identified numerous factors that align with all levels of influence in Carson et al.’s (2014) conceptual framework. These factors include public policies (e.g., state mandates related to whole-of-school PA; Phelps et al., 2018), school culture (e.g., prioritizing PA amid the pressures of academic testing, PA role-modelling by school staff, celebrating PA participation; Cassar et al., 2022; Ní Chróinín & McMullen; 2020), program leadership (e.g., a person who champions the initiative; McMullen et al., 2022), administrative support (e.g., providing teachers with PA promotion resources, allowing for teacher autonomy in program implementation, administrators role-modelling PA; Cassar et al., 2022; Phelps et al., 2018), facilities (e.g., physical education facilities, bike racks that support active commuting), (Phelps et al., 2018), knowledge and beliefs related to PA promotion (Cassar et al., 2022) and the availability of many PA opportunities (McMullen et al., 2022). Such research is critical to informing ongoing and future efforts to maximize the implementation of whole-of-school PA initiatives.

In parallel with global trends, 81% of children and adolescents in the United Arab Emirates are not accumulating at least 60 minutes of moderate-to-vigorous PA each day (Alrahma et al., 2023). One initiative that may help to address this issue is the Dubai Fitness Challenge. Since 2017, the Dubai government has implemented the Dubai Fitness Challenge (also referred to as Dubai 30×30) annually with the goal of engaging Dubai residents in 30 minutes of fitness activities for 30 days (late October through late November). Numerous activities are organized throughout the month, such as a half marathon, a bicycle ride, sports competitions, video workouts and free fitness classes. According to the Khaleej Times (citing data from Dubai’s Department of Economy and Tourism) there were 786,000 participants in the first year of the event’s implementation and 1.6 million participants in 2021 (close to half of Dubai’s total population) and 88% of those who participated in 2021 reached the 30/30 goal.

Though not a requirement, Dubai schools are strongly encouraged to participate in Dubai 30×30. Some media attention has been given to reporting various kinds of activities that schools implement during the initiative and the benefits of the initiative according to physical education teachers (“Dubai Fitness Challenge”, 2022; Roberts, 2022; “Schools Get Moving”, 2022). Examples of activities include partnering with community organizations/companies to offer different sports, games, and fitness events, taking field trips (e.g., to the beach) to exercise, having students use activity passports or scorecards to incentivize participation, and engaging school staff and parents in activity participation.

Reported benefits include students wanting to continue doing activities they were introduced to throughout the month and parents’ enjoying participation in the initiative. Despite these media portrayals of Dubai 30×30 implementation in schools, there is a lack of research that examines such implementation. No studies have investigated schools’ implementation of the initiative from the perspective of whole-of-school PA promotion, which is essential for helping school leaders (e.g., government officials who oversee the education sector in Dubai, school administrators) optimize Dubai 30×30 implementation using available school and community resources. The purpose of this study, therefore, was to examine schools’ implementation of Dubai 30×30 through a whole-of-school PA lens. As this was the first study to investigate schools’ implementation of Dubai 30×30, we opted to approach the study in a broad and exploratory manner by asking the following research questions: (a) What was the extent and nature of Dubai 30×30 activities for students, staff, and families before, during and after school? (b) To what extent did students participate in Dubai 30×30 activities? (c) How did students’ PA levels during school relate to contextual variables? (d) To what extent and in what ways were staff, families and community partners involved/engaged in promoting Dubai 30×30? (e) What were the perceived successes and challenges involved with implementing the initiative? and (f) What was the perceived impact of implementing the initiative on students, staff, families, and the community?

## Materials and Methods

### Research Design

This descriptive study was conducted using a convergent parallel mixed-methods research design (Edmonds & Kennedy, 2017). Specifically, quantitative and qualitative data were collected concurrently and analyzed jointly to address the research questions.

### Participants and Setting

Convenience sampling was used to recruit three private international schools in Dubai to participate in the study. Each school follows a British curriculum. The first school (School 1) serves students ages 3-11 and the second and third schools (School 2 and School 3) serve students ages 3-18. School 1 opened in 1989, School 2 opened in 2015 and School 3 opened in 2021. At the time of this study, more information was available for Schools 1 and 2 than for School 3 because the first two schools had undergone government inspections and reports of the inspection results were publicly available online, whereas the third school had not yet been inspected. School 1 had a total enrollment of 1177 students and a teacher-student ratio of 1:15. School 2 had a total enrollment of 1020 students and a teacher-student ratio of 1:8. School 3 had a total enrollment of 1290 students but there was no information available about teacher-student ratio. The largest nationality group of students at School 1 was UK, while the largest nationality group of students for School 2 was European. There was no information available about students’ nationalities for School 3. For the 2022-2023 school year, the Dubai government rated School 1 as “outstanding” and School 2 as “very good.” These ratings encompass numerous areas of focus, such as students’ achievement, personal and social development, teaching and assessment, staffing, facilities, and other resources. While School 3 had not yet received a rating, our visits to the school led us to believe that the learning climate and availability of resources were similar in quality compared to the first two schools.

Participant details from each school are displayed in Table 1. There were four participant groups: students, physical education teachers, classroom teachers and principals. Participation is reported in terms of the number of individuals from each participant group who participated in each method of data collection (survey, observations, and focus groups/interviews; see the following section on data sources and procedures). In all, there were 78 survey participants, 15 classes that were observed and 29 focus group/interview participants. Age bands for the different school years (grades) are 5-6 for Year 1, 6-7 for Year 2, 7-8 for Year 3, 8-9 for Year 4, 9-10 for Year 5, 10-11 for Year 6 and 11-12 for Year 7.

**Table 1.**
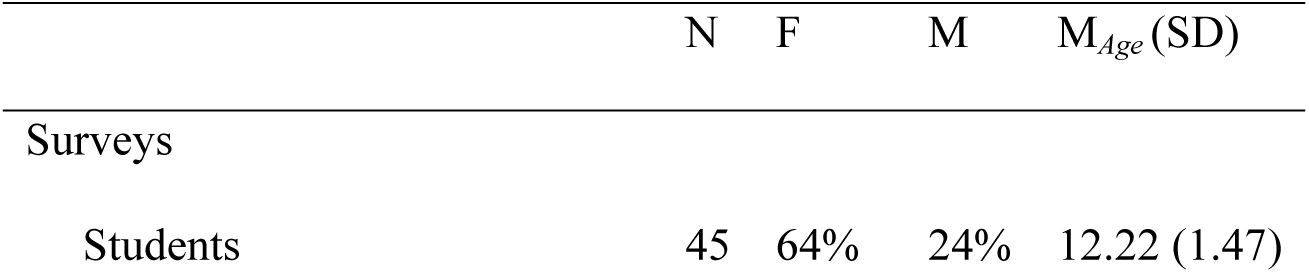
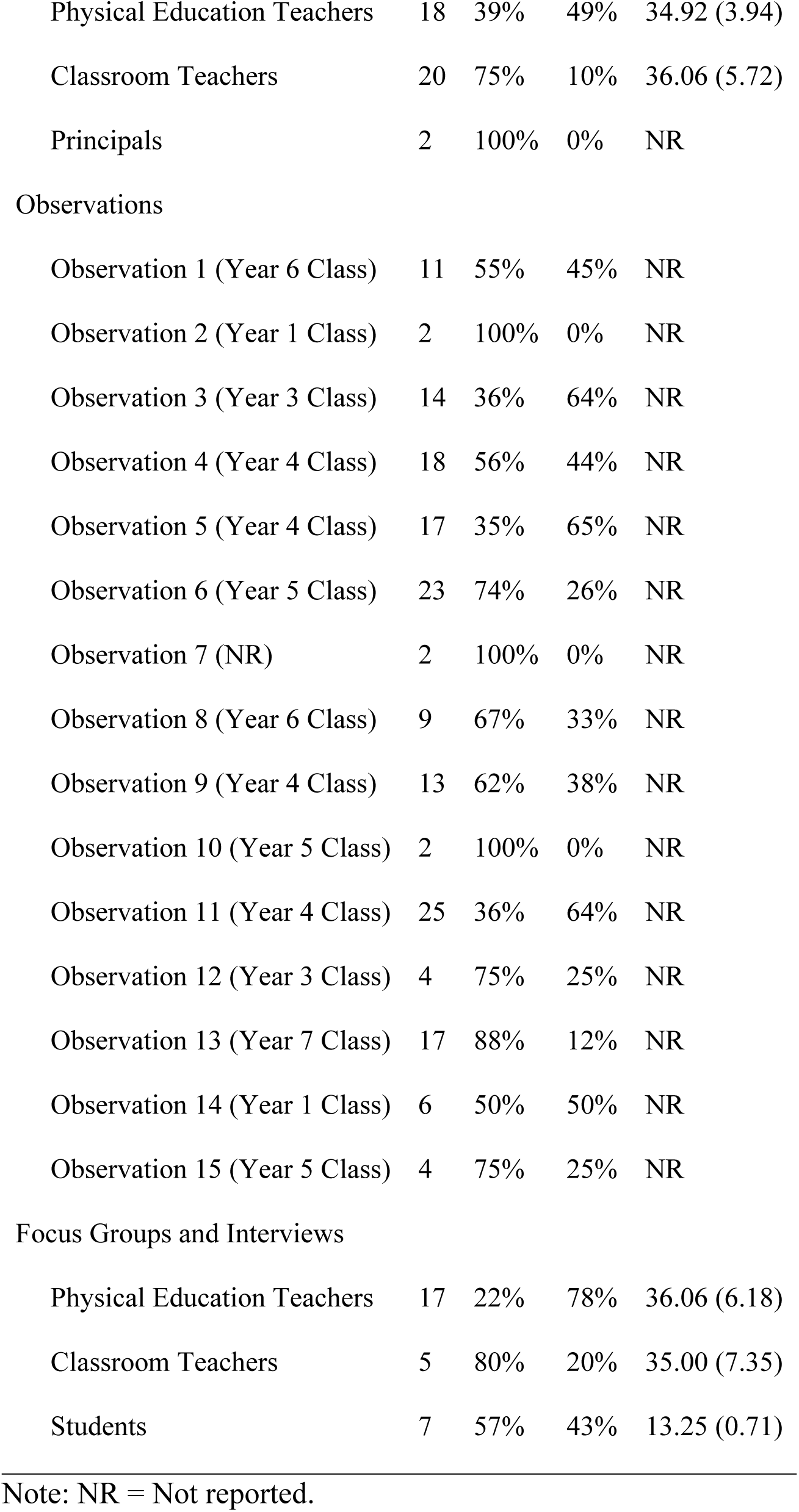
Participant Details.

*Participant Details*
|  | N | F | M | M<sub>Age</sub> (SD) |
| --- | --- | --- | --- | --- |
| Surveys |  |  |  |  |
| Students | 45 | 64% | 24% | 12.22 (1.47) |
| Physical Education Teachers | 18 | 39% | 49% | 34.92 (3.94) |
| Classroom Teachers | 20 | 75% | 10% | 36.06 (5.72) |
| Principals | 2 | 100% | 0% | NR |
| Observations |  |  |  |  |
| Observation 1 (Year 6 Class) | 11 | 55% | 45% | NR |
| Observation 2 (Year 1 Class) | 2 | 100% | 0% | NR |
| Observation 3 (Year 3 Class) | 14 | 36% | 64% | NR |
| Observation 4 (Year 4 Class) | 18 | 56% | 44% | NR |
| Observation 5 (Year 4 Class) | 17 | 35% | 65% | NR |
| Observation 6 (Year 5 Class) | 23 | 74% | 26% | NR |
| Observation 7 (NR) | 2 | 100% | 0% | NR |
| Observation 8 (Year 6 Class) | 9 | 67% | 33% | NR |
| Observation 9 (Year 4 Class) | 13 | 62% | 38% | NR |
| Observation 10 (Year 5 Class) | 2 | 100% | 0% | NR |
| Observation 11 (Year 4 Class) | 25 | 36% | 64% | NR |
| Observation 12 (Year 3 Class) | 4 | 75% | 25% | NR |
| Observation 13 (Year 7 Class) | 17 | 88% | 12% | NR |
| Observation 14 (Year 1 Class) | 6 | 50% | 50% | NR |
| Observation 15 (Year 5 Class) | 4 | 75% | 25% | NR |
| Focus Groups and Interviews |  |  |  |  |
| Physical Education Teachers | 17 | 22% | 78% | 36.06 (6.18) |
| Classroom Teachers | 5 | 80% | 20% | 35.00 (7.35) |
| Students | 7 | 57% | 43% | 13.25 (0.71) |

Note: NR = Not reported.

### Data Sources and Procedures

The Ethics Board at the University of Birmingham approved this study. School and participant recruitment was conducted from August 8^th^, 2022 to January 30^th^, 2023.

Participants provided written informed consent prior to the start of data collection. Parents/guardians provided consent for student participants. Data sources for this study included surveys, school observations, focus groups and individual interviews. The surveys and school observations provided quantitative data, while the focus groups and individual interviews provided qualitative data. A physical education teacher at each school served as the point of contact during the study and helped to recruit participants and facilitate data collection.

#### Surveys

Schools 2 and 3 participated in electronic surveys that were administered over a three-week period directly following Dubai 30×30 (School 1 opted not to participate in the surveys). The online survey platform JISC was used to create and distribute the surveys and manage the data. Weekly reminders were sent via email to increase the number of survey respondents. There were four surveys: one for students, one for physical education teachers, one for classroom teachers and one for principals. Students completed their survey during normal classroom time and with assistance from teachers, as needed.

Survey items pertinent to the current study focused on Dubai 30×30 activities before, during and after school (Research Question 1), students’ participation in these activities (Research Question 2) and school staff’s involvement in Dubai 30×30 (Research Question 4). Regarding Dubai 30×30 activities, school staff were asked to indicate the extent to which activities were provided in each of six contexts (physical education, classroom, recess/break, before school, after school, and weekends), whereas students were asked to indicate their level of participation in these activities. A six-point response scale was used. The response scale for school professionals ranged from “No Dubai 30×30 activities were provided” to “A lot of Dubai 30×30 activities were provided.” Participants could also choose an option that stated, “I don’t know where there were any Dubai 30×30 activities provided.” The response scale for students ranged from “I did not participate at all in Dubai 30×30 activities” to “I participated a lot in Dubai 30×30 activities.” Students could also choose the option, “There were no Dubai 30×30 activities provided.”

Items assessing staff involvement were adapted from existing measures developed in previous whole-of-school research (Orendorff et al., 2021; Stoepker et al., 2021; Webster et al., 2013). Example items include “I am involved with Dubai 30×30 planning at my school” (asked to all school staff), “I am involved with integrating Dubai 30×30 activities into physical education lessons” (asked only to physical education teachers), “I am involved with integrating Dubai 30×30 activities into my classroom lessons or as breaks/transitions during normal classroom time” (asked only to classroom teachers) and “I am involved with allocating resources for my school’s participation in Dubai 30×30” (asked only to principals). A six-point response scale was used, which ranged from “Strongly Disagree” to “Strongly Agree.”

The surveys also included a demographic section at the end. All participants were asked to identify their gender and their race/ethnicity. Additionally, students were asked to identify their age, while school staff were asked to indicate the levels at which they were teaching (e.g., elementary, secondary). An optional response format was used for all items on the surveys so that participants could skip questions if they wished to do so.

#### Observations

The Observational System for Recording Physical Activity in Children – Elementary School (OSRAC-E; McIver et al., 2016) was used to examine the relationship between students’ PA levels and contextual variables during school (Research Question 3).

Although the instrument was designed for elementary school settings, much of the category system can be applied to secondary school settings, as well. Five categories from the instrument were used for the purposes of the present study: activity level, location, physical setting, instructional setting, and activity context. OSRAC-E uses a focal child protocol, whereby a single child is observed at a time using momentary time sampling. We employed a five-second observe interval followed by a 25-second record interval, which was repeated for five minutes with the same child. This observation procedure was used with as many children as possible during each school visit during the study.

In the week before observations were scheduled to commence, the second author conducted a two-hour training session on the observation protocols used for the study.

Trainees were undergraduate students who volunteered to help with data collection and other aspects of the research. The session involved familiarizing the trainees with the OSRAC-E and testing the trainees’ reliability in using the instrument. Inter-observer reliabilities were calculated using the following formula (van der Mars, 1989):

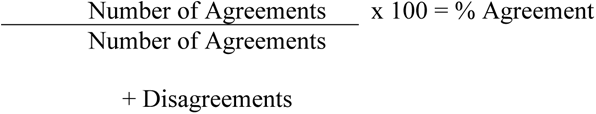

Video examples were used for the training and to test inter-observer reliability. A reliability standard of 80% was considered acceptable (van der Mars, 1989). Total of eight trainees demonstrated acceptable inter-observer agreement proceeded with conducting observations the following week.

A second two-hour training session was held after the first week of data collection to help prevent observer drift and address any unanticipated challenges with data collection. During this session, observers shared their experiences doing the observations and suggested ways to help increase inter-observer reliability. Suggestions mainly focused on how to record unusual activities or types of behavior using the OSRAC-E. The session also included additional opportunities for observers to practice using the using more video examples and concluded with another round of reliability testing, which resulted in acceptable levels of agreement (above 80%) for all observers.

Observations were conducted across 15 classrooms from Schools 2 and 3 during the full month of Dubai 30×30 (School 1 opted not to participate in the observations).

Observation days and times were scheduled in accordance with when the observers were available and when the schools were able to accommodate observers. In total, observations were conducted for 10.25 hours across 12 days. During each school visit, observers usually stayed with a single class of students and followed the class during the full observation period (approximately 2-7 hours). This allowed for data to be collected in different instructional contexts (e.g., homeroom, physical education). Observation shifts lasted two and a half hours for each observer to help prevent exhaustion and loss of focus. Additional inter-observer reliability checks were conducted in the third and fourth weeks of data collection with agreement scores indicating acceptable reliability.

#### Focus groups and interviews

A total of six focus groups and two individual interviews were conducted (see Table 1 for participant details). The purpose of the focus groups and interviews was two-fold. First, they were intended to provide additional information about Dubai 30×30 activities (Research Question 1) and the involvement/engagement of school staff, families, and community partners in promoting Dubai 30×30 (Research Question 4). Second, they were designed to address school staff’s perceived successes and challenges involved with implementing Dubai 30×30 (Research Question 5) and participants’ perceived impact of the initiative on students, families, and the community (Research Question 6). A semi-structured format was used to allow for some flexibility in each focus group/interview. Examples of questions include “What did your school do in the name of Dubai 30×30 this school year?”, “Describe your involvement in Dubai 30×30 this school year as a physical education/classroom teacher/principal. What roles did you play?”, “From your perspective, what were notable successes for your school during its involvement with Dubai 30×30 this school year?”, “Describe any challenges or problems you remember from your school’s involvement with Dubai 30×30 this school year?” and “What was the impact of your school’s involvement in Dubai 30×30?” Additionally, prompts were used for clarification and to explore participants’ responses in more depth. Following recommended protocols, two researchers conducted each focus group with one researcher asking questions and the other researcher taking notes and helping to moderate, when needed (Krueger & Casey, 2014). Interviews/focus groups generally lasted 30 minutes, were conducted either in person at participants’ schools or online via Zoom and were audio recorded.

### Data Analysis

#### Surveys

Microsoft Excel was used to calculate descriptive statistics, including means and standard deviations, for the extent of Dubai 30×30 activities before, during and after school (Research Question 1), the extent of students’ participation in Dubai 30×30 activities (Research Question 2) and the extent of involvement/engagement of school staff in promoting Dubai 30×30 (Research Question 4).

#### Observations

To examine the relationship between students’ PA and contextual variables during school (Research Question 3), the likelihood of students engaging in each activity level was estimated using multinomial logistic regression with students engaging in stationary as the referent group. Analyses were completed using STATA (v.16.1, STATA Corporation LLC, College Station, TX).

#### Focus groups and interviews

All audio recordings were transcribed verbatim for analysis. Open and axial coding were used to develop themes in the data (Strauss & Corbin, 1990). Specifically, the first author read each transcript to become familiar with participants’ responses. Then, he initiated open coding, which involved writing memos that summarized/interpreted the responses (or parts of responses) in each transcript that seemed helpful in addressing Research Questions 1, 4, 5 and 6. This process continued iteratively, allowing for revisions and additions to the memos as the author made additional passes through the transcripts and developed his understanding of the major ideas underlying the data. Subsequently, the author used axial coding to search across transcripts for memos suggesting similar ideas. He synthesized these memos into broader ideas related to each research question to develop themes. To help ensure trustworthiness (Lincoln & Guba, 1985), the second author served as a peer debriefer throughout the analysis, regularly meeting with the first author to discuss the analysis process, the coding, and the themes. Trustworthiness was also established through the triangulation of multiple data sources.

## Results

### Research Question 1: Dubai 30×30 Activities Before, During and After School

Descriptive statistics and themes related to Research Question 1 are shown in Table 2. School staff indicated on their surveys that there were more Dubai 30×30 activities provided during physical education than in other contexts. Recess/other breaks during school and before and after school periods were also identified as times when there were a fair amount of Dubai 30×30 activities provided. Participants felt that there were very few Dubai 30×30 activities provided during classroom time and on weekends. Themes and supporting evidence from the focus group and interview data are presented in Table 3. The themes converged with the survey data in that all participant groups (physical education teachers, classroom teachers and students) and the principal spoke about Dubai 30×30 activities that were offered during break times and before and after school. There was also a theme characterized by a recurring competitive element to many of the activities. Surprisingly, physical education was seldom mentioned in relation to Dubai 30×30 activities, which contrasted with the survey data.

**Table 2.**
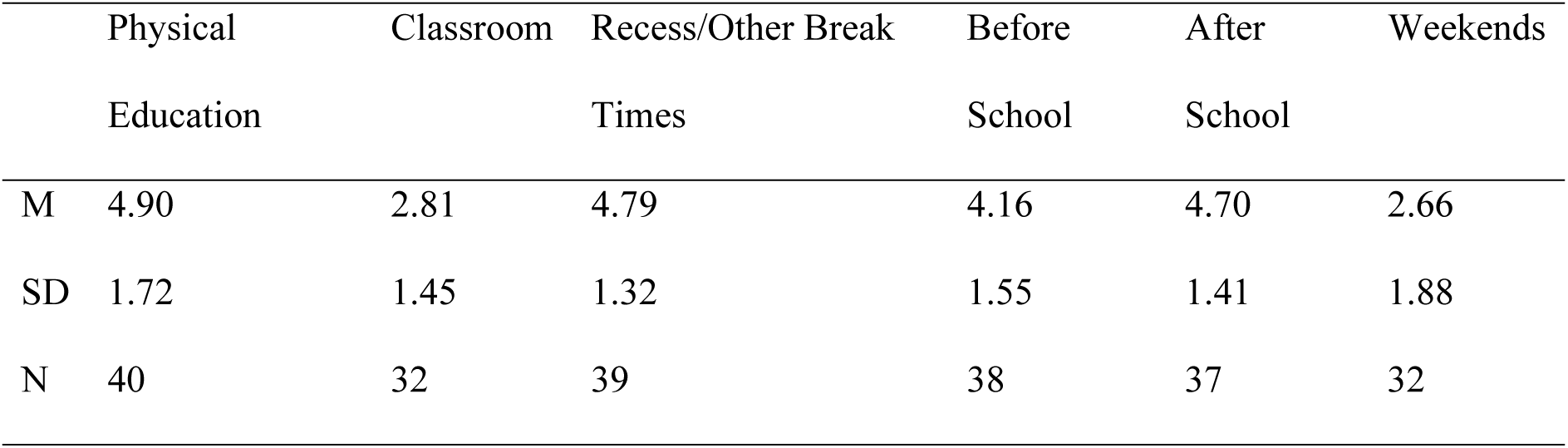
Research Question 1: Extent of Dubai 30×30 Activities Provided Before, During and After School (Combined Descriptive Data from Surveys Administered to Physical Education Teachers, Classroom Teachers, and Principals)

**Table 3.**
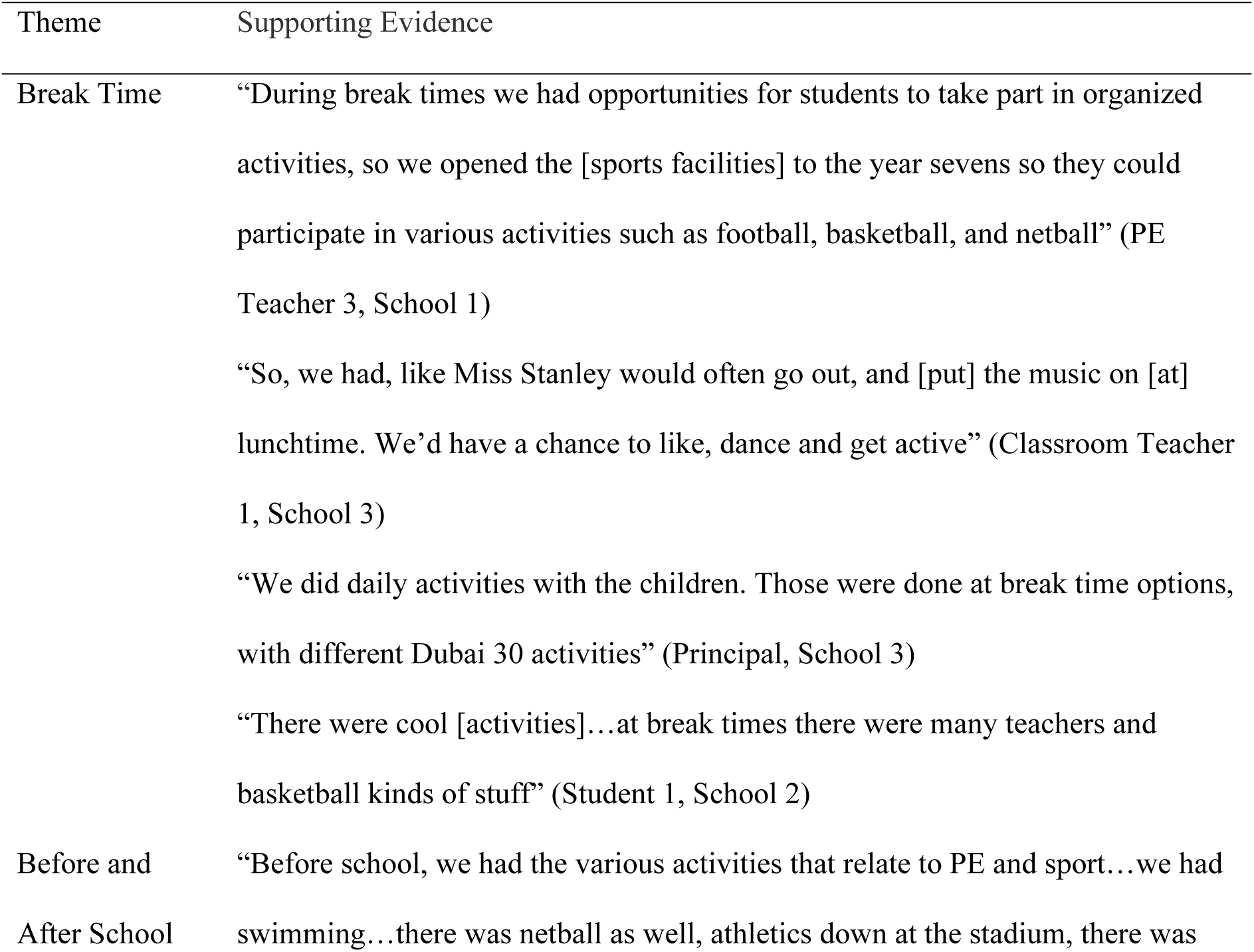
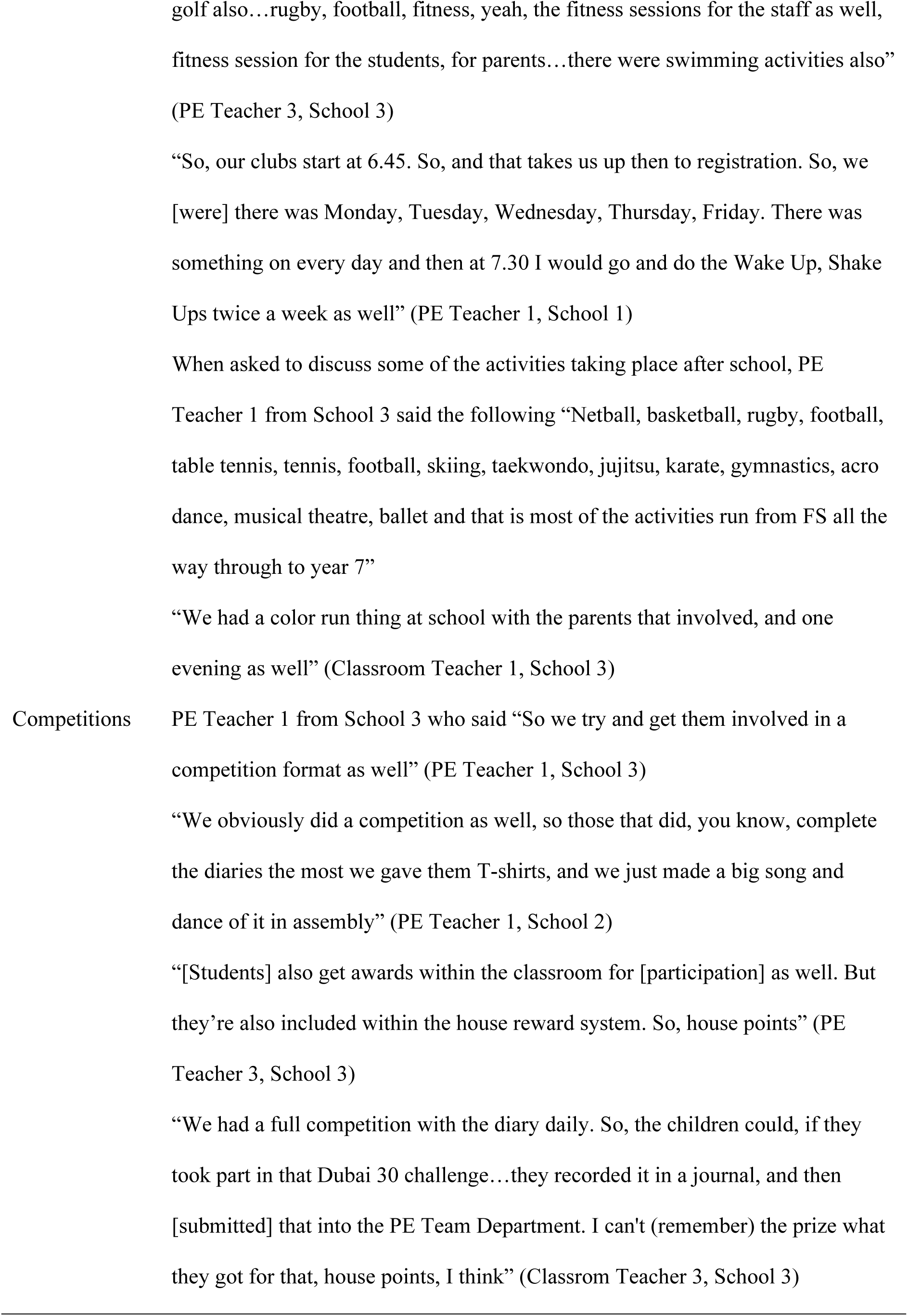
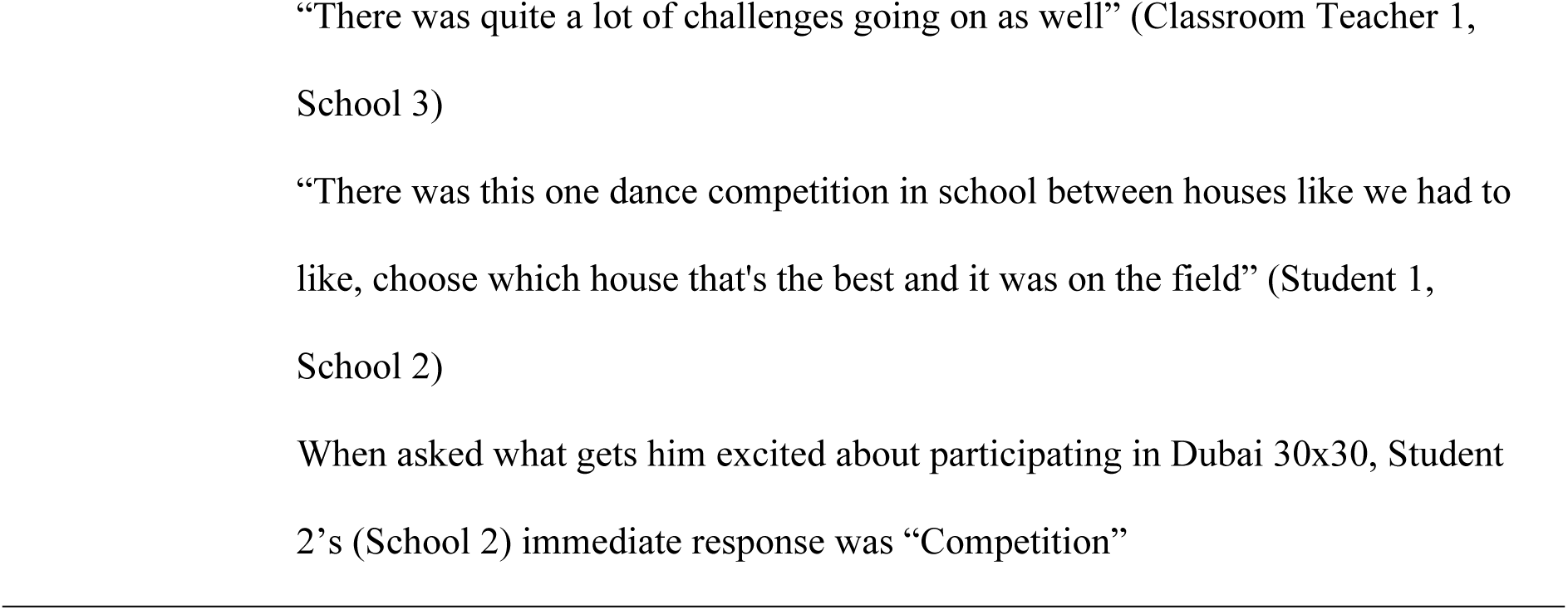
Research Question 1: Extent and Nature of Dubai 30×30 Activities Provided Before, During and After School (Themes and Supporting Evidence from Focus Group and Interview Data)

### Research Question 2: Students’ Participation in Dubai 30×30 Activities

Table 4 displays the descriptive statistics related to Research Question 2. Consistent with school staff’s survey responses regarding the extent of Dubai 30×30 activities provided in different contexts, students reported participating a fair amount in Dubai 30×30 activities during physical education and very little in such activities during classroom time. However, in contrast to the school staff survey data, students indicated they participated very little in Dubai 30×30 activities during recess/other break times and occasionally participated in initiative-related activities after school and on weekends.

**Table 4.** Research Question 2: Extent of Students’ Participation in Dubai 30×30 Activities Before, During and After School (Data from Survey Administered to Students)

|  | Physical Education | Classroom | Recess/Other Break Times | Before School | After School | Weekends |
| --- | --- | --- | --- | --- | --- | --- |
| M | 4.20 | 2.79 | 2.70 | 2.24 | 3.0 | 3.19 |
| SD | 1.56 | 1.61 | 1.70 | 1.67 | 1.77 | 1.85 |
| N | 45 | 38 | 40 | 41 | 44 | 44 |

### Research Question 3: Relationship between Students’ PA Levels and Contextual Variables During School

Results of the multinomial logistic regression models are presented in Table 5. Several physical setting variables were related to students’ engagement in activity. For example, children were 2.39 (95CI=1.08, 5.28) and 52.36 (95CI=13.12, >100.00) times as likely to engage in slow easy and fast activity in the gym compared to the classroom.

**Table 5.**
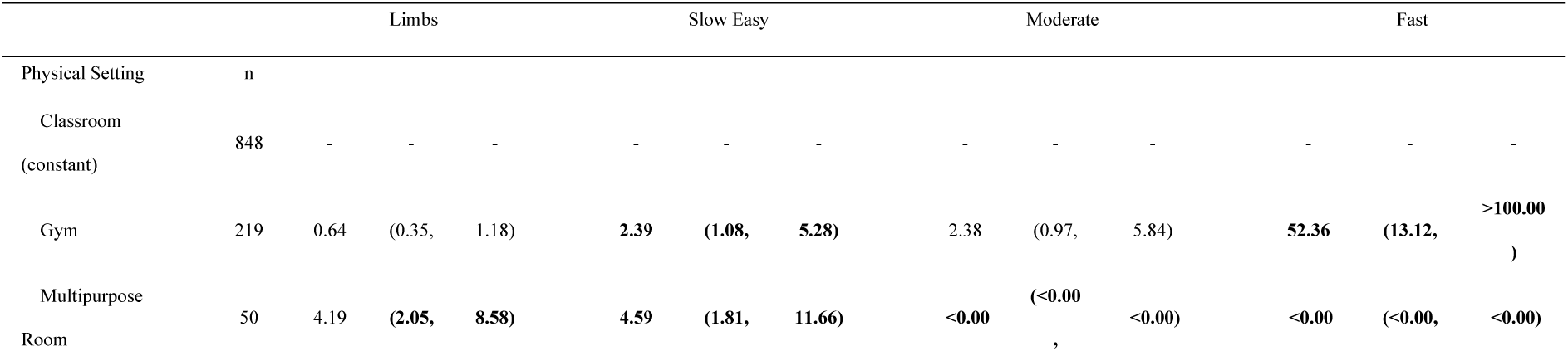
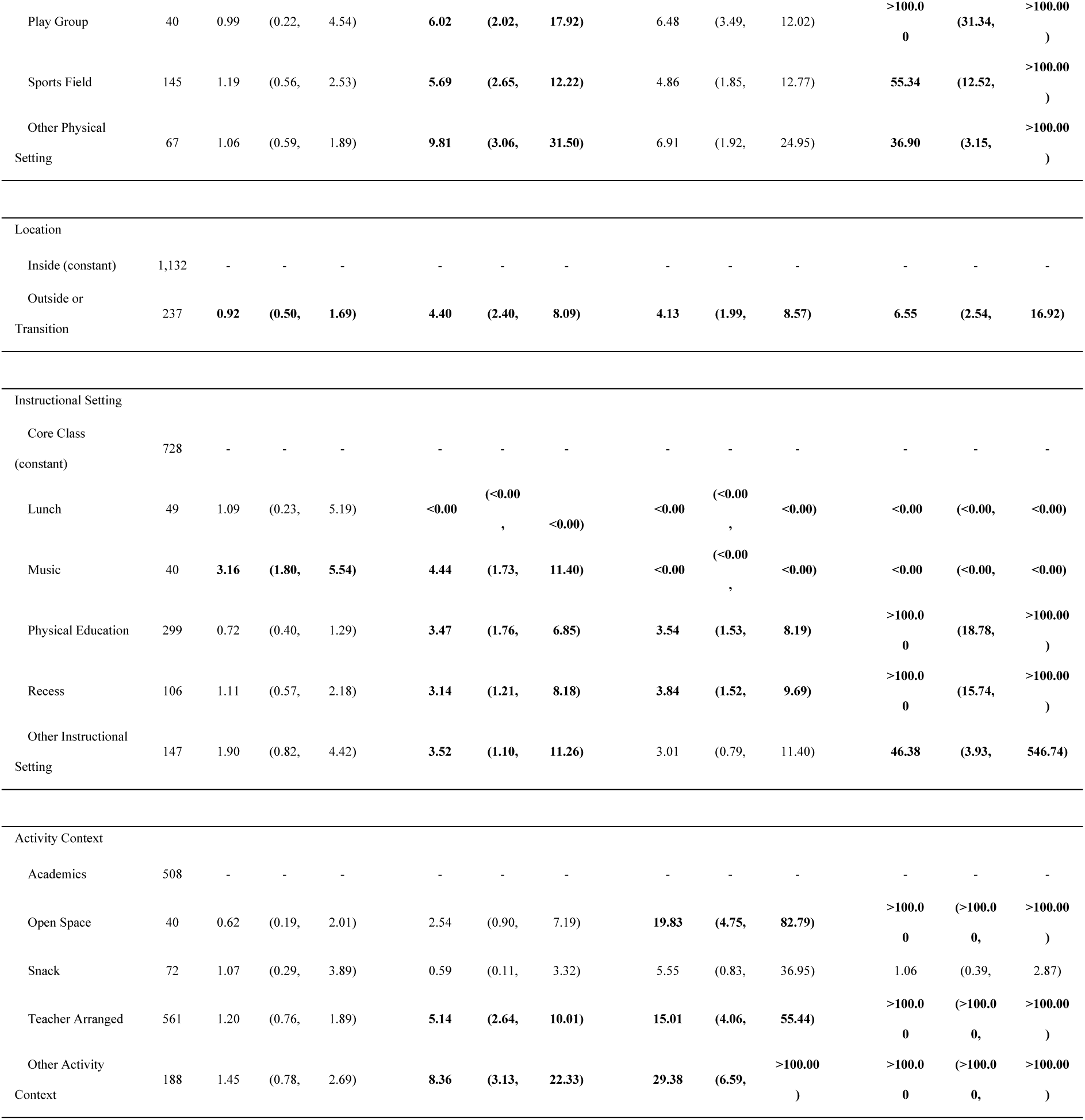
Research Question 3: Relationship between Students’ PA Levels and School Context (Multinomial Logistic Regression of Activity Levels by OSRAC-E Variables)

Further, the playground, sports fields and other physical settings showed similar relationships with children’s physical activity. At the playground children were 6.02 (95CI=2.02, 17.92), 6.48 (95CI=3.49, 12.02) and 110.66 (95CI=31.34, >100.00) times as likely to engage in slow easy, moderate, and fast activity compared to the classroom. At the sports field children were 5.69 (95CI=2.65, 12.02), 4.86 (95CI=1.85, 12.77) and 55.34 (95CI=12.52, >100.00) times as likely to engage in slow easy, moderate, and fast activity compared to the classroom. In other physical settings children were 9.81 (95CI=3.06, 31.50), 6.91 (95CI=1.92, 24.95) and 36.90 (95CI=3.15, >100.00) times as likely to engage in slow easy, moderate, and fast activity compared to the classroom. In the multipurpose room children were 4.19 (95CI=2.05, 8.58) and 4.59 (95CI=1.81, 11.66) times as likely to engage in limbs and slow easy, but <0.00 (95CI= <0.00, <0.00) times as likely to engage in both moderate and fast activity when compared to the classroom.

Location, instructional setting and activity context were also related to children‘s activity level. Children were 4.40 (95CI=2.40, 8.09), 4.13 (95CI=1.99, 8.57), and 6.55 (95CI=2.54, 16.92) times as likely to engage in slow easy, moderate, and fast activity when outside or in transition compared to when they were inside. With respect to instructional setting, children were <0.00 (95CI= <0.00, <0.00) times as likely to engage in slow easy, moderate, or fast activity during lunch compared to core class time. Conversely, during music, children were 3.16 (95CI=1.80, 5.54) and 4.44 (95CI=1.73, 11.40) times as likely to engage in limbs and slow easy activity, but <0.00 (95CI= <0.00, <0.00) times as likely to engage in moderate or fast activity when compared to core class time. During physical education, children were 3.47 (95CI=1.76, 6.85), 3.54 (95CI=1.53, 8.19), and >100.00 (95CI=18.78, >100.00) times as likely to engage in slow easy, moderate, and fast activity compared to core class time. Similarly, during recess children were 3.14 (95CI=1.21, 8.18), 3.84 (95CI=1.52, 9.69), and >100.00 (95CI=15.74, >100.00) times as likely to engage in slow easy, moderate, and fast activity compared to core class time. In other instructional settings, children were 3.52 (95CI=1.10, 11.26) and 46.38 (95CI=3.93, >100.00) s likely to engage in slow easy and fast activity compared to core class time.

Regarding activity context, children were 19.83 (95CI=4.75, 82.79), and >100.00 (95CI=>100.00, >100.00) times as likely to engage in moderate and fast activity during open space activities compared to academic activities. During teacher organized activities, children were 5.14 (95CI=2.64, 10.01), 15.01 (95CI=4.06, 55.44), and >100.00 (95CI=>100.00, >100.00) times as likely to engage in slow easy, moderate, and fast activity compared to academic activities. Finally, during other activity contexts children were 8.36 (95CI=3.13, 22.33), 29.38 (95CI=6.59, >100.00), and >100.00 (95CI=>100.00, >100.00) times as likely to engage in slow easy, moderate, and fast activity compared to academic activities.

### Research Question 4: Involvement/Engagement of School Staff, Families and Community Partners

Table 6 displays the descriptive statistics related to Research Question 4. Overall, physical education teachers’ perceived involvement in promoting Dubai 30×30 was higher than classroom teachers’ or principals’ perceived involvement. Physical education teachers somewhat agreed that they were involved in integrating Dubai 30×30 activities into their lessons, promoting Dubai 30×30 activities outside of physical education, generating support from others to implement Dubai 30×30 activities, finding solutions to problems with implementing Dubai 30×30 activities, planning Dubai 30×30 activities, organizing Dubai 30×30 activities, advocating for the school’s involvement in Dubai 30×30 and serving as a health/fitness role model. Principals’ perceived involvement was second highest overall in comparison to physical education teachers and classroom teachers. Principals agreed that they were involved with allocating resources for their school’s participation in Dubai 30×30 and advocating for their school’s involvement in Dubai 30×30. They somewhat agreed that they were involved in evaluating the school’s participation in Dubai 30×30, establishing Dubai 30×30 activities at the school, setting performance standards for the school’s participation in Dubai 30×30, building/maintaining partnerships for implementing/sustaining Dubai 30×30, planning for Dubai 30×30 implementation and being a healthy/fit role model. Classroom teachers disagreed/somewhat disagreed about their involvement across all relevant items.

**Table 6.**
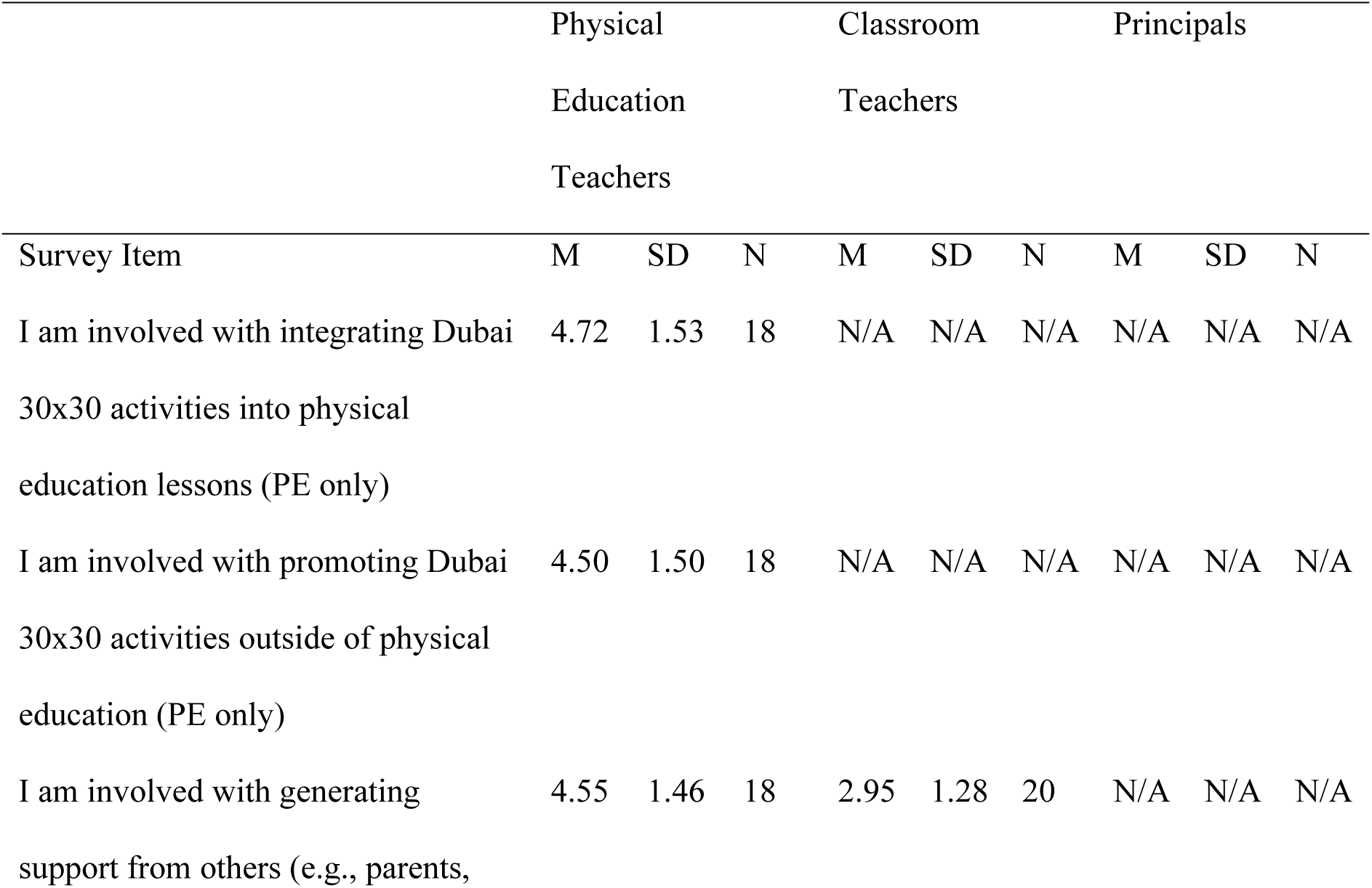
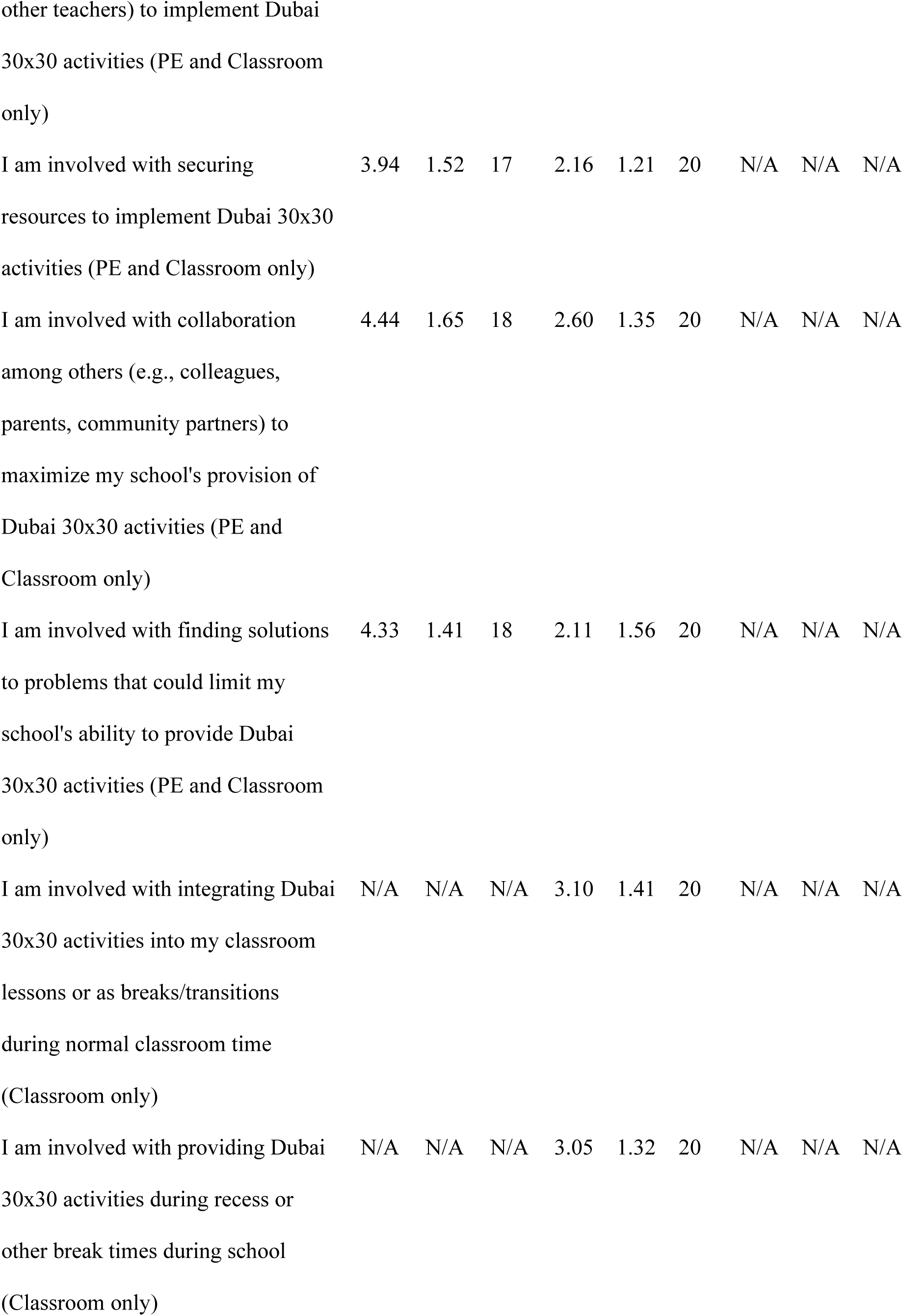
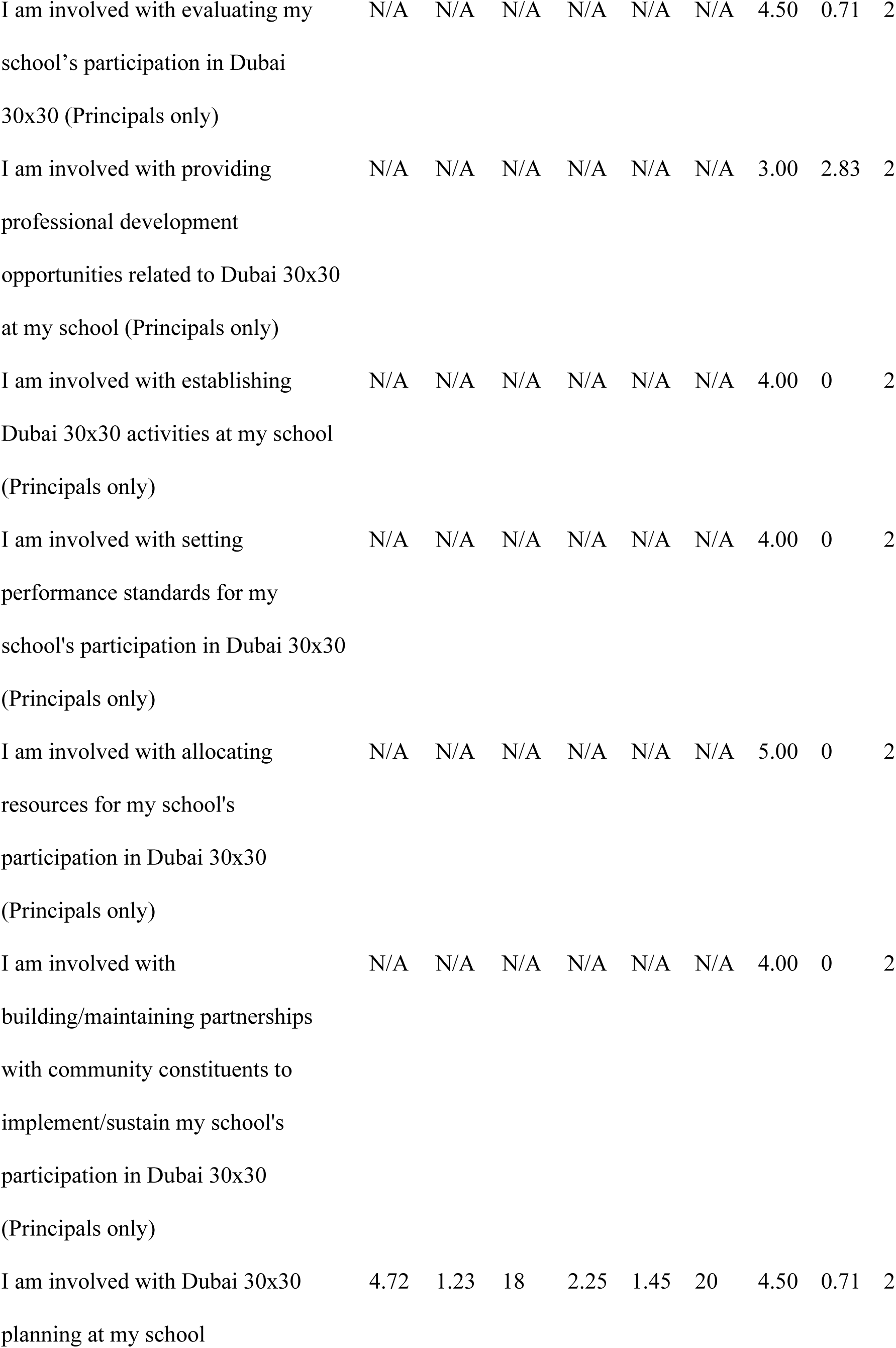
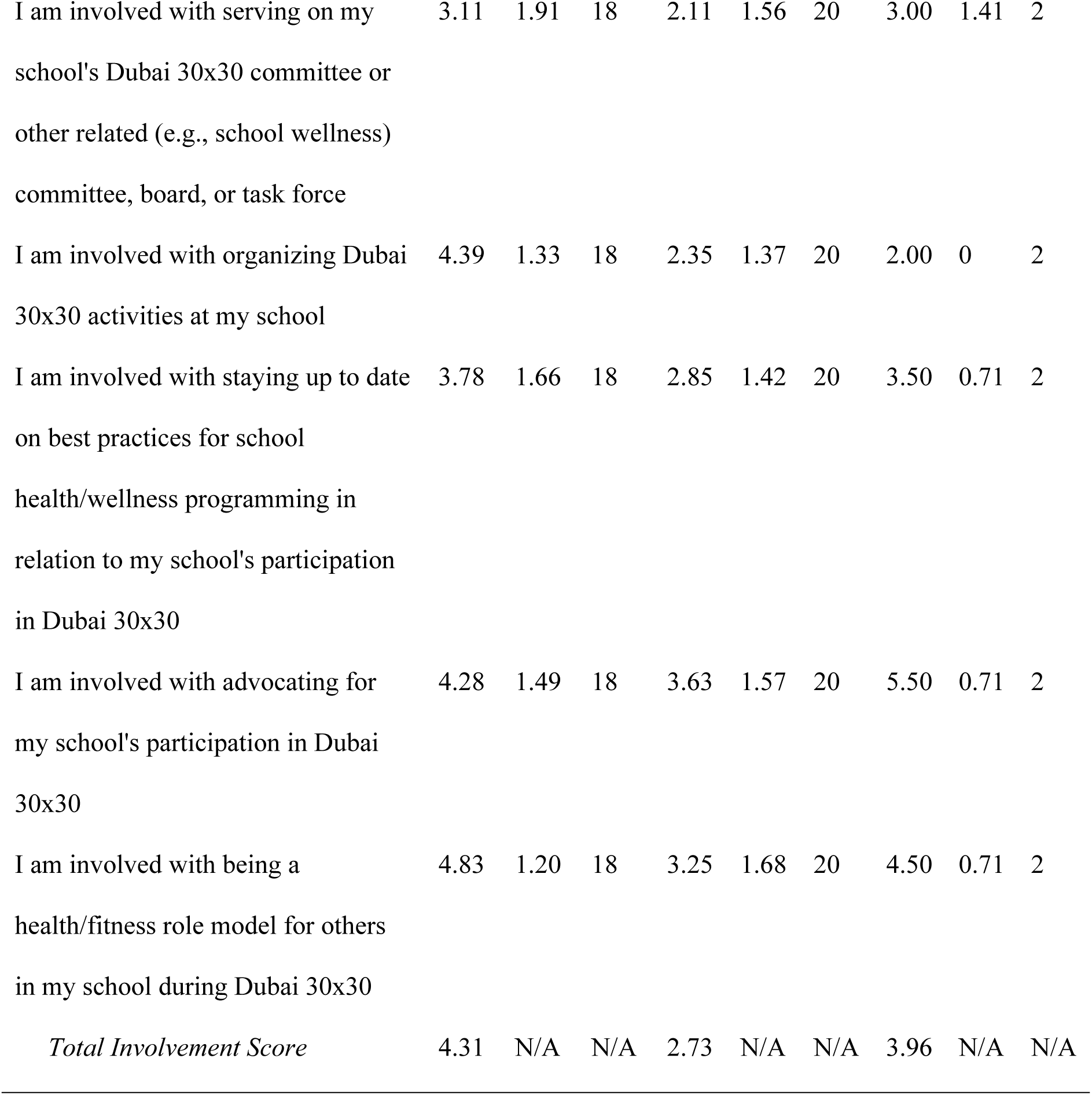
Research Question 4: Involvement/Engagement of School Staff in Promoting Dubai 30×30 (Data from Surveys Administered to Physical Education Teachers, Classroom Teachers and Principals)

Themes and supporting evidence related to Research Question 4 are presented in Table 7. The first theme, “Physical Education Teachers as Dubai 30×30 Leaders,” closely aligns with the survey data, further demonstrating that physical education teachers were distinctly involved in schools’ implementation of the initiative. The common approach was for someone on the physical education team to spearhead the planning and organization of Dubai 30×30 activities, guide the other physical education teachers in their efforts to support the implementation of the activities and serve as the primary liaison for other school staff, parents and any other entities who were involved. The second them focused on parent engagement, which mainly entailed parents participating in various activities that the school provided, some of which were specifically designed for parents.

**Table 7.**
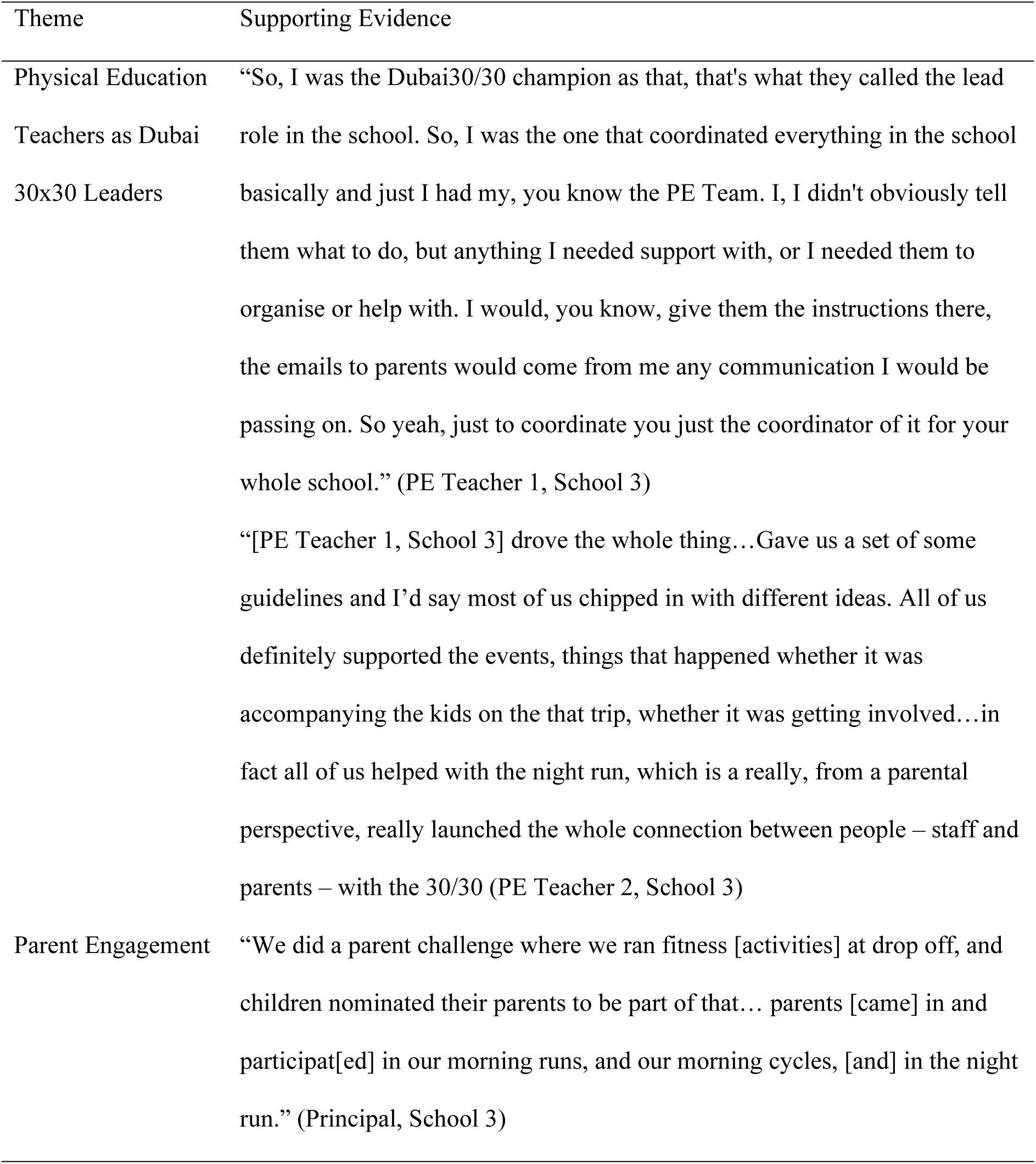
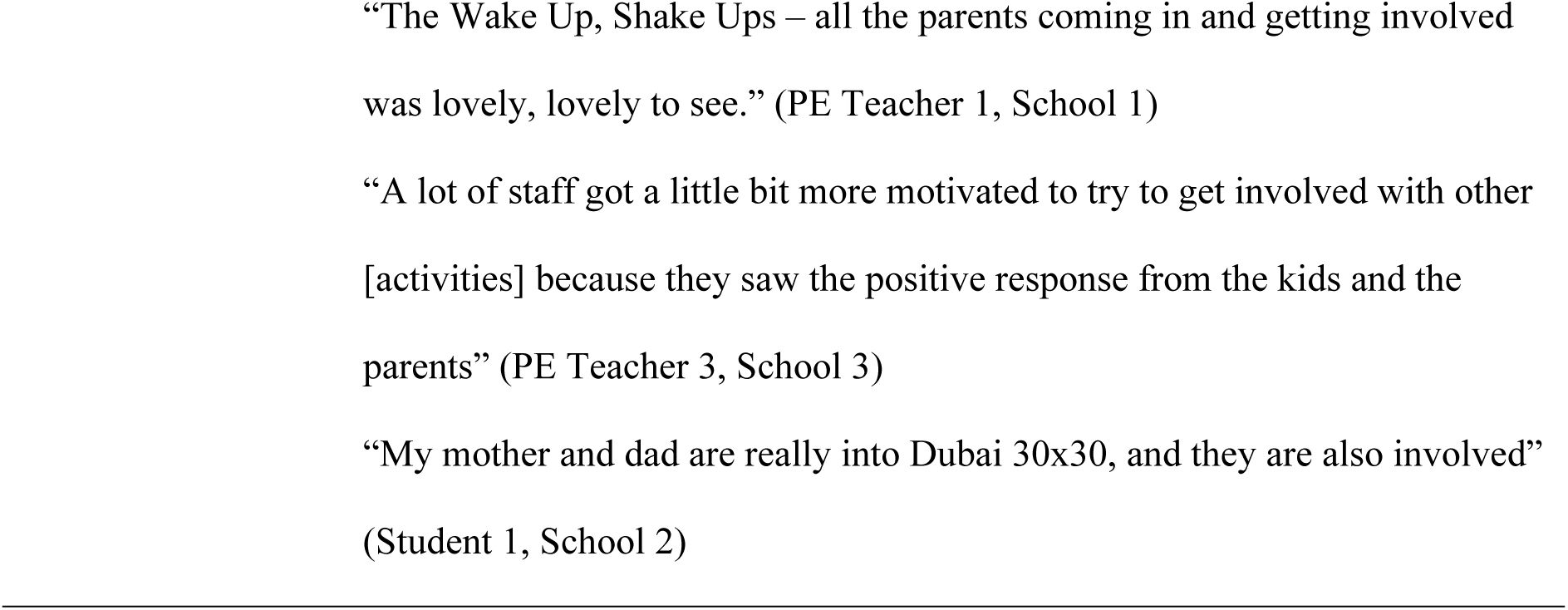
Research Question 4: Involvement/Engagement of School Staff, Families and Community Partners in Promoting Dubai 30×30 (Themes and Supporting Evidence from School Staff Focus Group and Interview Data)

### Research Question 5: School Staff’s Perceived Successes and Challenges of Dubai 30×30 Implementation

Themes and supporting evidence related to Research Question 5 are presented in Table 8. Implementation successes were described in terms of communities coming together. Implementation challenges were mainly discussed relative to the lack of guidance and the pressures placed on physical education staff.

**Table 8.**
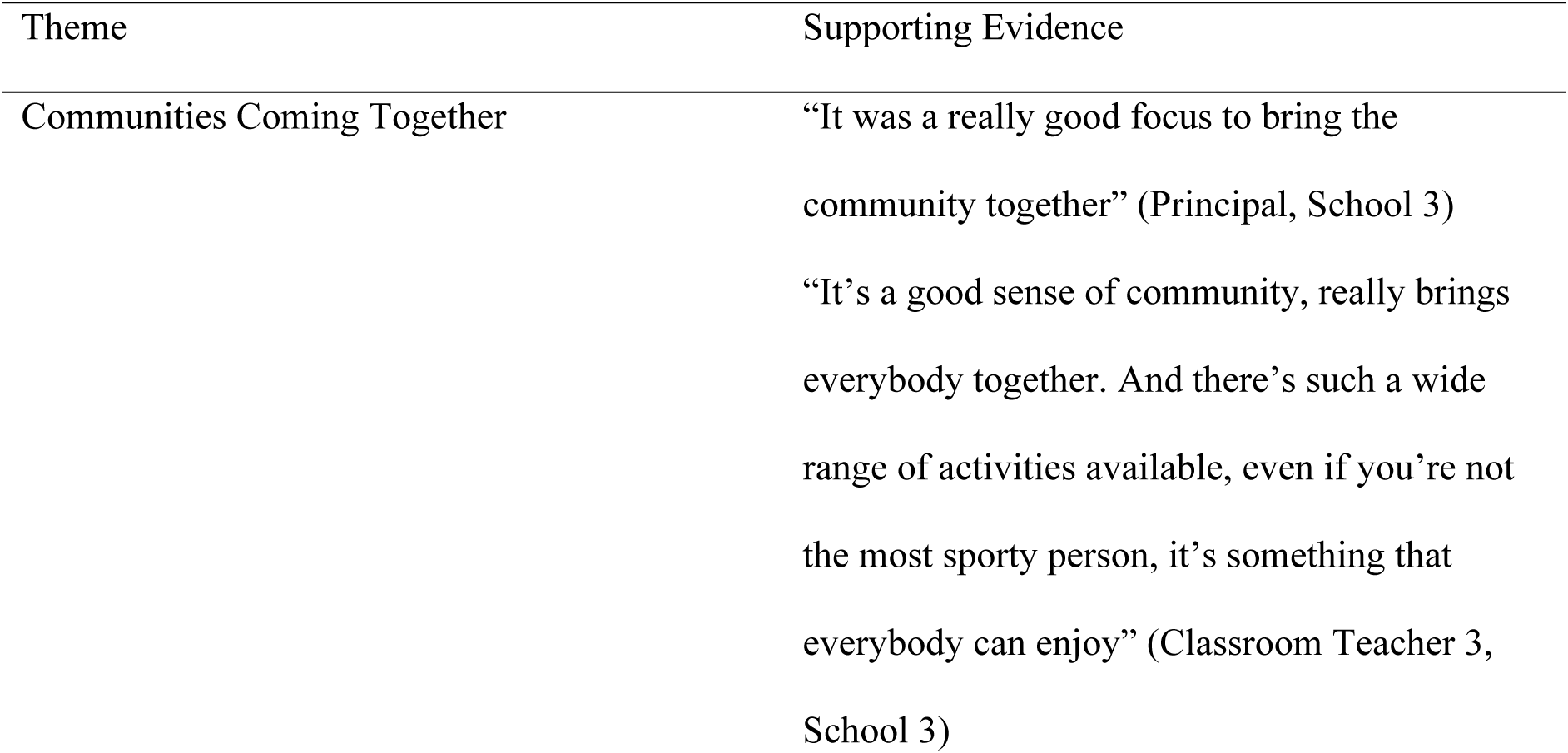
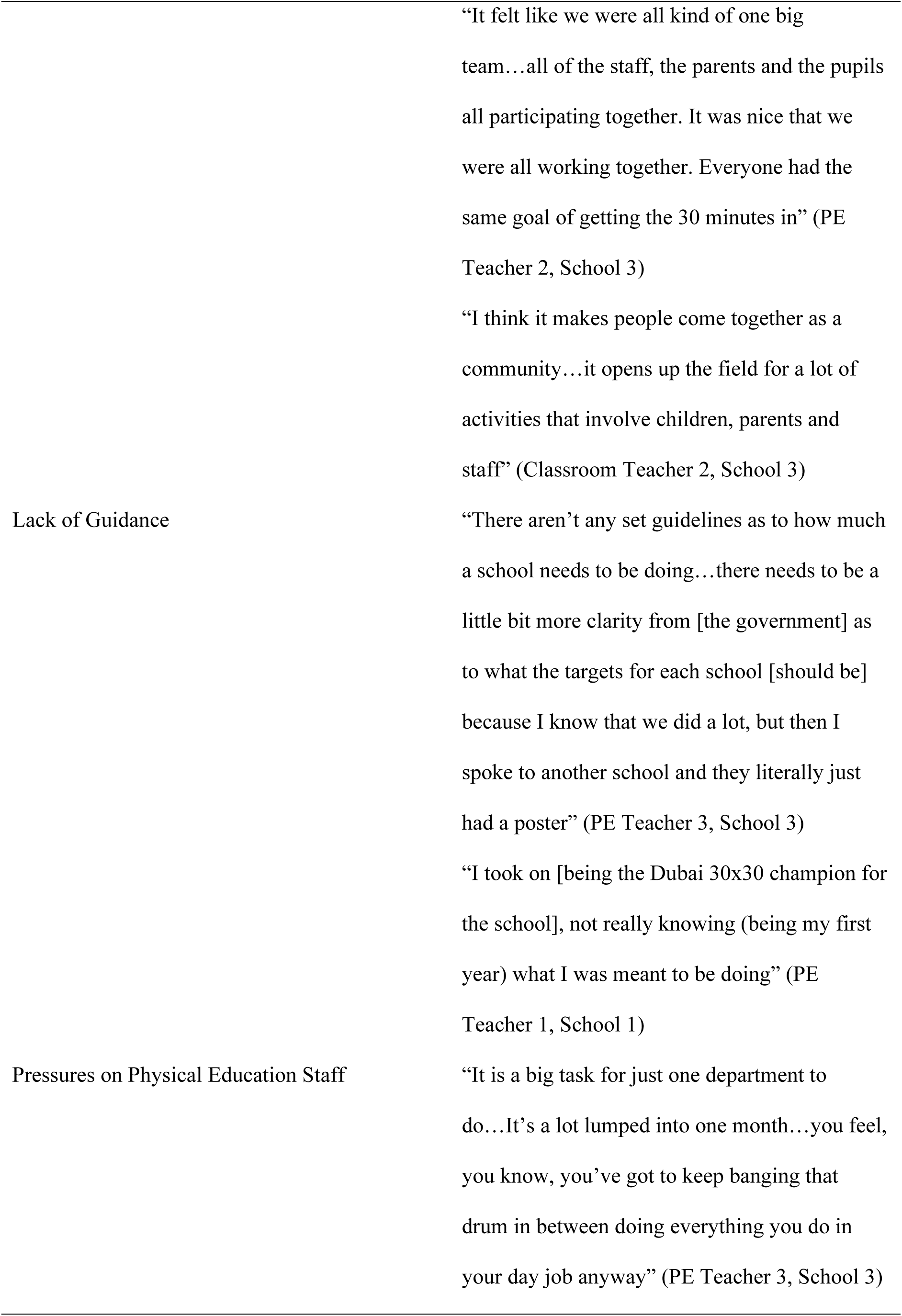
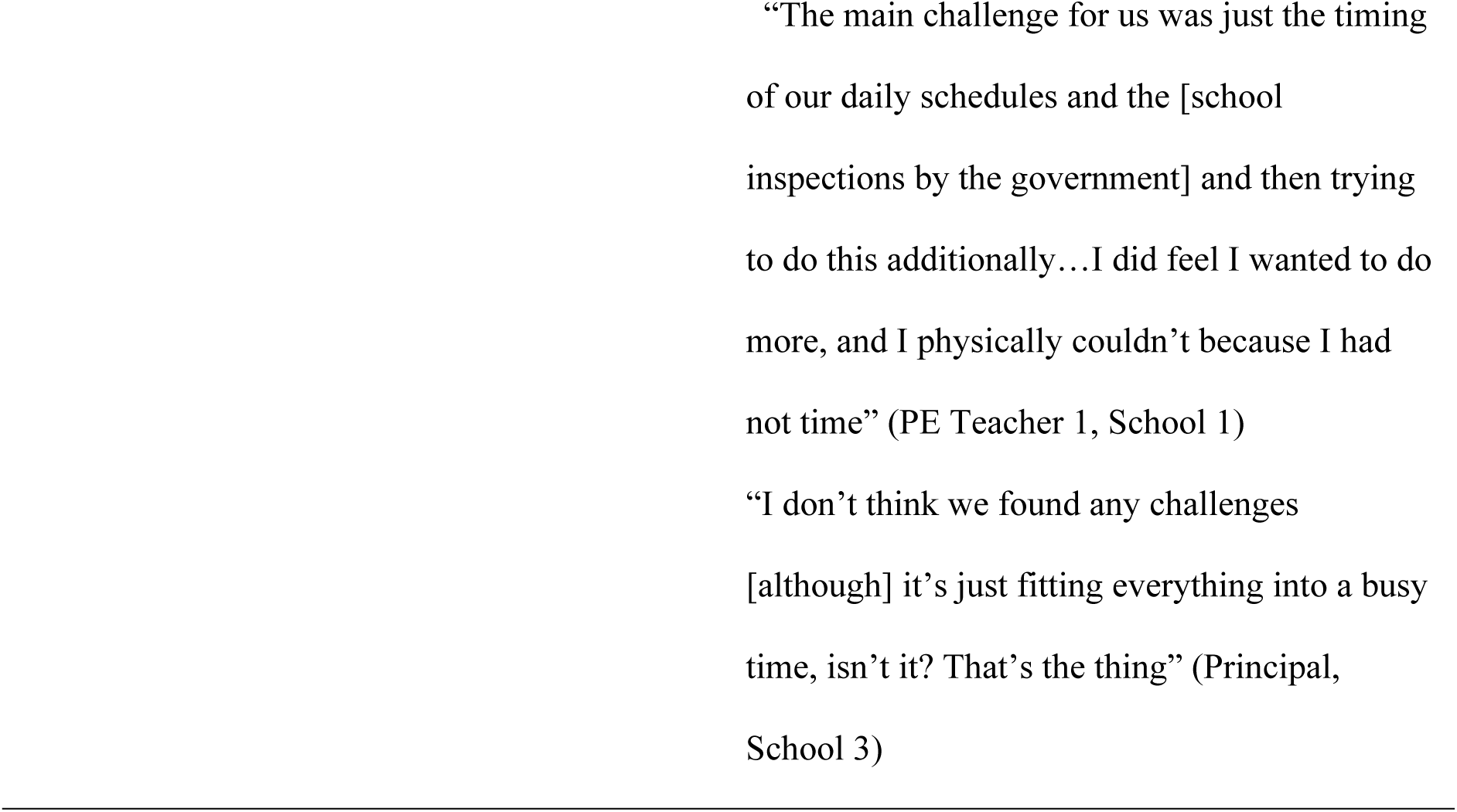
Research Question 5: School Staff’s Perceived Implementation Successes and Challenges (Themes and Supporting Evidence from School Staff Focus Group and Interview Data)

### Research Question 6: Participants’ Perceived Impact of Dubai 30×30

The themes and supporting evidence related to Research Question 6 are displayed in Table 9. There were two themes: (a) increased PA and (b) promotion of the school. Many of the statements participants made about the impact of Dubai 30×30 focused on increased PA for youth and parents. Promotion of the school constituted a smaller theme that became apparent within the transcripts from participants at School 3.

**Table 9.**
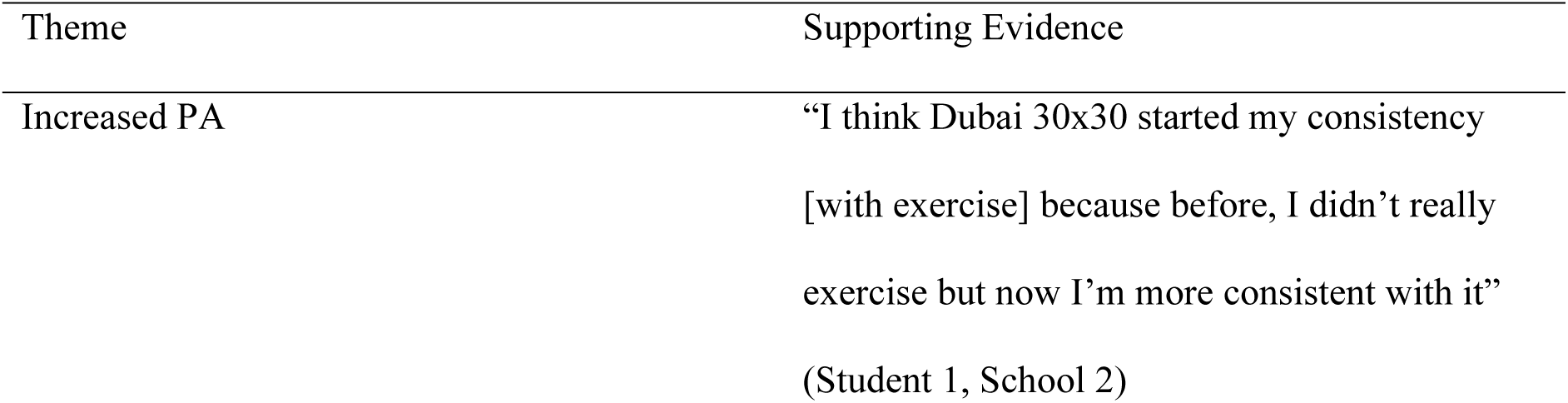
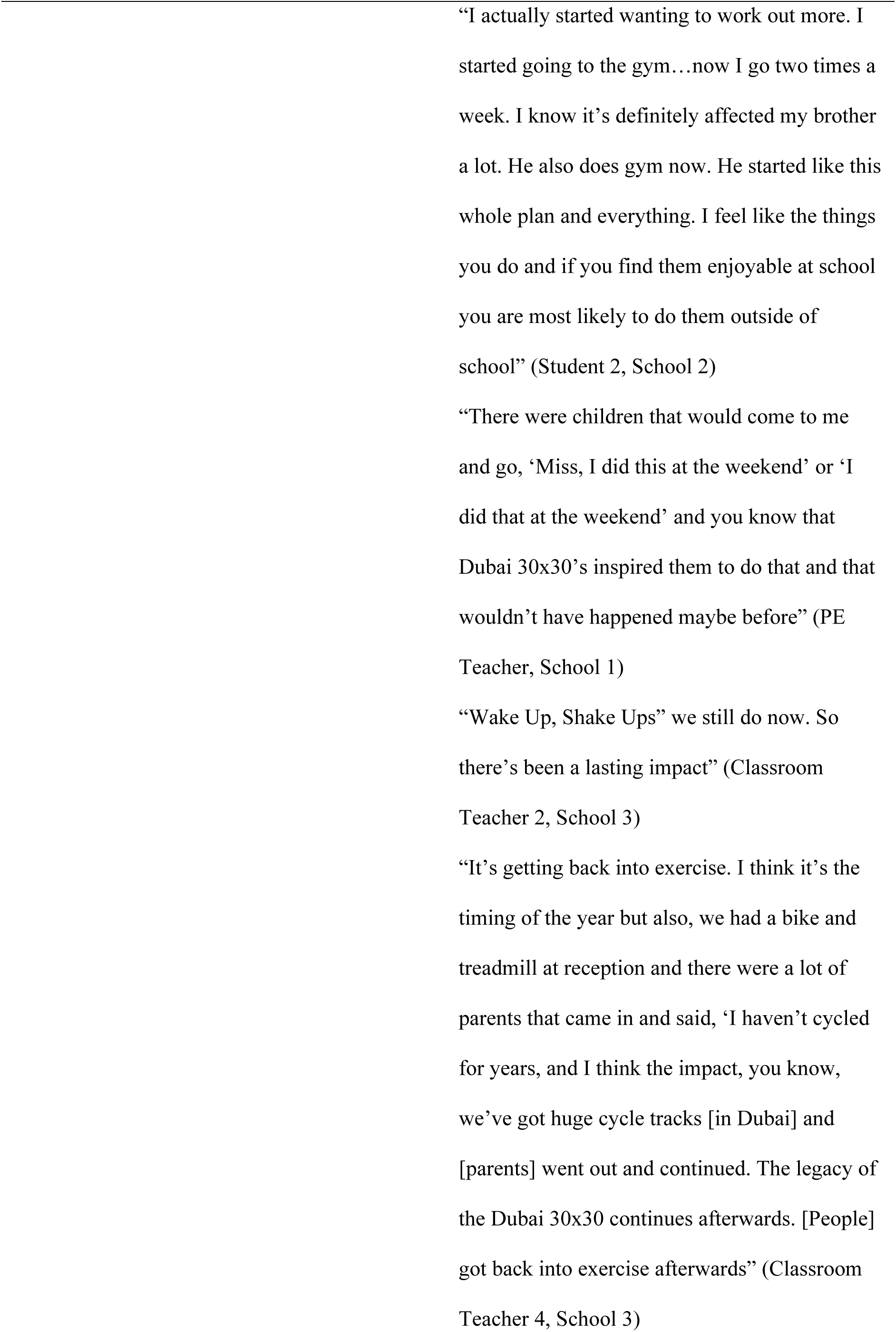
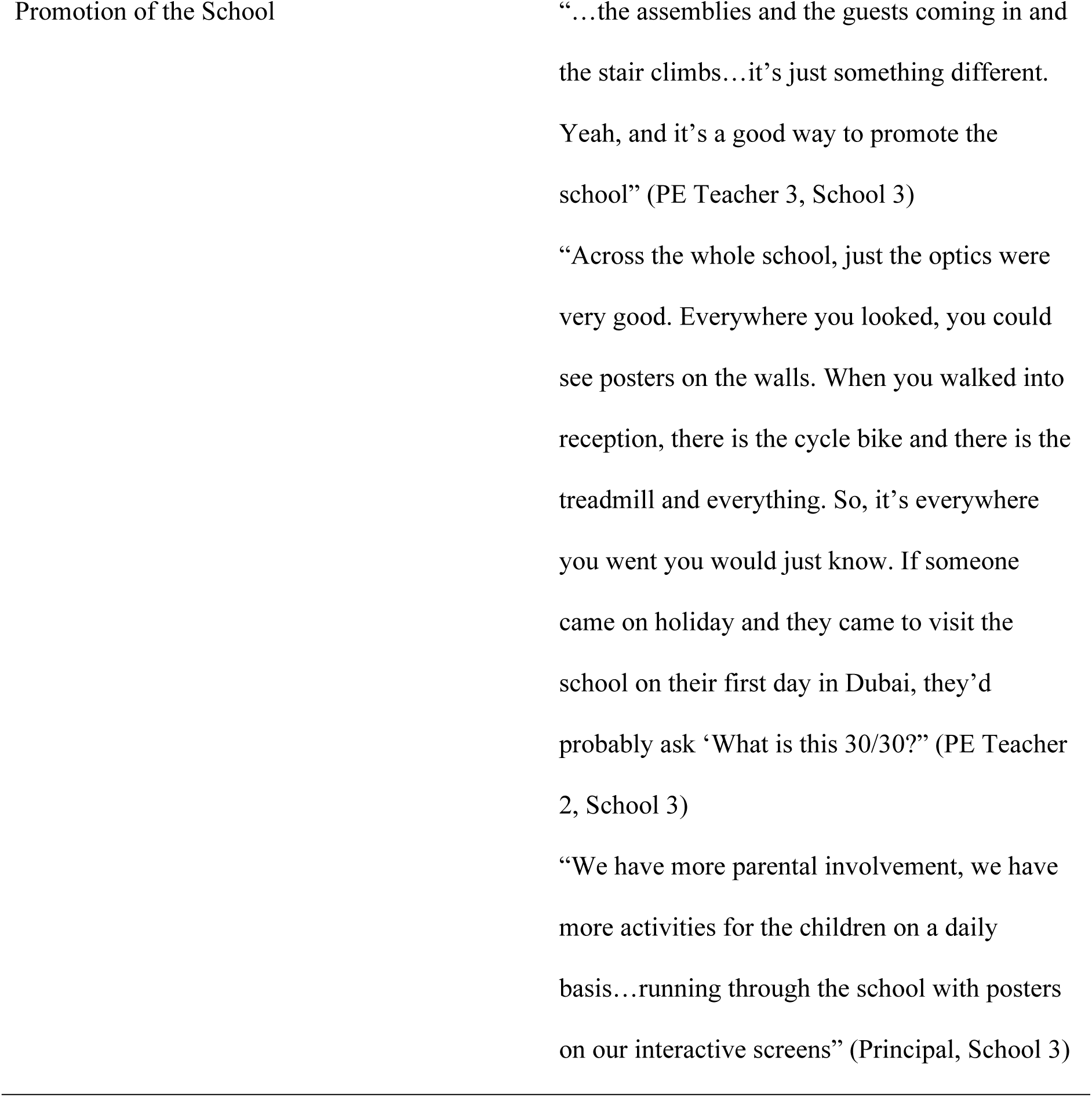
Research Question 6: Perceived Impact of Dubai 30×30 (Themes and Supporting Evidence from Focus Group and Interview Data)

## Discussion

Consistent with global trends in PA among school-age youth (Guthold et al., 2020), most children and adolescents in the UAE are insufficiently active (Alrahma et al., 2023). The current study drew upon conceptions of whole-of-school PA promotion (CDC, 2019; IOM, 2013; Milton et al., 2021) to examine schools’ participation in Dubai 30×30. To our knowledge, this is the first study to provide descriptive data about implementation and outcomes related to schools’ involvement with Dubai 30×30. We therefore adopted an exploratory approach, driven by several research questions intended to provide an initial glimpse into various aspects of whole-of-school PA. Specifically, this study investigated the scope and nature of Dubai 30×30 activities offered before, during and after school and on weekends; students’ participation in Dubai 30×30 activities; the role of school contextual variables in students’ PA during Dubai 30×30; staff involvement in promoting Dubai 30×30; staff’s perceptions of implementation successes and challenges; and staff’s and students’ perceptions of the impact of Dubai 30×30. The results of this study contribute to the growing knowledge base internationally on whole-of-school PA approaches, point to potential directions for future research and establish preliminary evidence that can be used to optimize Dubai 30×30 as a lever for increasing and sustaining whole-of-school PA in Dubai schools.

### Research Question 1: Dubai 30×30 Activities Provided

School staff reported that most Dubai 30×30 activities were provided in physical education, during school break times and before and after school. This suggests that Dubai 30×30 activities spanned many of the contexts identified in whole-of-school PA recommendations (CDC, 2019; IOM, 2013; Milton et al., 2021). Internationally, there are inconsistencies in the degree to which schools provide PA through all possible recommended components/practices of a whole-of-school approach (American Alliance for Health, Physical Education, Recreation and Dance, 2011; Brenner et al., 2017; Colabianchi, et al., 2015; WHO, 2022). For example, in a national study of U.S. secondary schools, 7% of middle school students and less than 1% of high school students reported attending schools that offered all six practices conceptualized as a whole-of-school approach (having physical education; offering PA breaks during the school day; having intramural sports; having interscholastic sports; having active transportation opportunities to/from school; and having shared-use of school facilities agreements with outside entities; (Colabianchi et al., 2015).

The results of a meta-analysis study indicated that increasing the number of components through which school-based PA programming is provided is positively associated with an increase in students’ total daily PA minutes (Russ et al., 2015). However, Webster et al. (2020) assert that school-based PA programming should be viewed as sufficient based on whether all students achieve the goals of the program, as opposed to whether the program consists of a certain number of components or practices. Future research on schools’ implementation of Dubai 30×30 should further investigate both the quantity and quality of PA offerings and the association of such offerings with targeted student outcomes (e.g., meeting PA guidelines, meeting academic standards).

### Research Question 2: Students’ Participation in Dubai 30×30 Activities

Students reported that they mainly participated in Dubai 30×30 activities during physical education and occasionally participated in activities after school and on weekends but infrequently participated in activities offered during break times. Comparing these data to the results for the first research question, it seems that providing opportunities for PA during break times may not always lead to students participating in such opportunities. This might have to do with students’ motivation. Lonsdale et al. (2009) found that adolescents in Hong Kong who were more self-determined in their motivation to participate in physical education had higher PA levels during a free-choice period compared to students who were less self-determined in their motivation. Therefore, a possible direction for future research on schools’ participation in Dubai 30×30 is to examine the relationship between students’ motivation and their participation in Dubai 30×30 activities. Additionally, the qualities or characteristics of Dubai 30×30 activities that students find most appealing, and which serve as sources of motivation to participate warrant increased attention in future studies. The present study revealed that students were drawn to competitive activities. Other whole-of-school research also highlighted competition as a key driver for students’ involvement in PA opportunities, although recognition of participation and effort was also important for fostering involvement (Ní Chróinín et al. 2020). In another study, some adolescents’ PA levels were highest when non-competitive physical activities were provided as part of a comprehensive approach to school health (Pirrie et al., 2022).

### Research Question 3: Relationship of Students’ PA to School Contextual Variables

During school, students were more likely to reach higher PA intensity levels when they were in locations, instructional settings, and activity contexts other than the regular classroom setting, core class time and academic activities. These findings reinforce the data addressing the first two research questions in that the participating schools appeared to place a limited focus on Dubai 30×30 as a catalyst or incubator for increasing PA during regular classroom time. Providing PA opportunities during periods of the day devoted to academic instruction (outside of physical education) – a practice referred to as movement integration (Webster et al., 2015) – is recommended as part of a whole-of-school PA approach (CDC, 2019; IOM, 2013; Milton et al., 2021). The current literature on movement integration mostly centers on primary/elementary school settings. In such contexts, movement integration in can involve various strategies, such as infusing PA into academic lessons or giving students brief breaks from sitting (Moon & Webster, 2019). Previous research shows that integrating PA during regular classroom time increases children’s PA participation and educational outcomes (Norris et al., 2020). However, classroom teachers often report barriers to using movement integration (e.g., lack of time, lack of resources, lack of support from school administrators; Michael et al., 2019; Schmidt et al., 2022). It would be interesting in future research to further explore the potential of using regular classroom time during Dubai 30×30 to increase children and adolescents’ school-based PA.

### Research Question 4: Involvement/Engagement of Staff, Families and Community Partners in Dubai 30×30

Among school staff, physical education teachers were most involved and classroom teachers were least involved in promoting Dubai 30×30, which resonates with the results for the first three research questions. Whole-of-school PA recommendations commonly call upon physical education teachers to serve as leaders in providing PA opportunities and galvanizing the support of other school staff, parents, and community partners (Beighle et al., 2009; Carson et al., 2014; McKenzie & Lounsbery, 2013). Yet, Webster et al. (2015) note that in many cases, certain obstacles may need to be overcome for physical education teachers to effectively lead whole-of-school PA initiatives. Such obstacles include a lack of professional preparation and training specific to whole-of-school PA leadership and a lack of incentives for physical education teachers to take on leadership responsibilities (e.g., personal interest, tangible rewards, external accountability). The findings in the present study seem to suggest that physical education teachers were successful in garnering involvement from parents and engaging numerous community partners. Principals also provided support for schools’ participation in Dubai 30×30. Thus, a priority for future implementations of the initiative will be to increase attention to helping physical education teachers build enhanced support from classroom teachers.

### Research Question 5: Staff’s Perceived Implementation Successes and Challenges

Staff perceived that Dubai 30×30 brought their school communities together. This is a fundamental goal of whole-of-school PA approaches, which are intended to draw upon the collaboration and synergy of school staff, families, and communities (CDC, 2019; IOM, 2013; Milton et al., 2021). However, physical education teachers also commented on the lack of guidance concerning implementation and they felt burdened by the amount of work involved and pressure to make Dubai 30×30 a special month for their schools. Unfortunately, we neglected to more fully explore the source of the pressure physical education teachers felt in relation to implementing Dubai 30×30 activities. In previous research, factors at multiple social-ecological levels exerted an influence (e.g., public policy, administrative support, teacher beliefs) on whole-of-school PA practices (Cassar et al., 2022; Phelps et al., 2018). In the case of Dubai 30×30, which the Dubai government initiated and strongly promotes, it is possible that the pressure physical education teachers feel may stem from the government, but it is also possible that other factors, such as the overall school ethos, principals’ priorities, parents’ interest, or physical education teachers’ own personal values contribute to these feelings of pressure.

Although principals noted that time was a barrier to implementation, neither they nor the classroom teachers communicated feelings of being overwhelmed, in contrast to what was evidenced in the focus groups and interview with physical education teachers. A more balanced diffusion of responsibility across all school stakeholders should be sought in future school-based implementations of the Dubai 30×30. For instance, consistent with recommendations for implementing whole-of-school PA programming (e.g., CDC, 2013), schools could form a school wide Dubai 30×30 committee to promote distributed leadership, shared decision-making, and clear guidance with respect to the initiative. Such a committee could a designated Dubai 30×30 “champion” (e.g., a physical education teacher), classroom teacher representatives, a school administrator, parent representatives, student representatives and community partner representatives.

### Research Question 6: Perceived Impact of Schools’ Implementation of Dubai 30×30

Participants believed Dubai 30×30 increased PA participation and helped to promote their schools. The value of these beliefs in energizing school staff and sustaining their involvement in the initiative is an important area of focus for future research. A study of school principals in the U.S. found that participants’ outcome expectations directly predicted their level of self-reported involvement in CSPAPs (Orendorff et al., 2020). Specifically, principals were more likely to be involved if they believed a CSPAP would benefit students not just in terms of their PA participation but also in terms of their physical development, cognitive development, social development, school attendance and academic performance.

Participants in the present study made little reference to most of these outcomes when discussing the impact of Dubai 30×30. In the future, researchers should more systematically measure a broad range of student outcomes associated with schools’ participation in Dubai 30×30, as such data may spur staff’s involvement.

### Limitations

This study had several limitations. First, although working with only a few schools allowed us to explore school-based Dubai 30×30 implementation from multiple perspectives, the use of a small convenience sample limits the generalizability of the results. Second, we sought to survey parents in this study, but no parents participated. Understanding the perspectives of parents and community partners is critical to advancing research and practice related to schools’ implementation of Dubai 30×30. Third, this study merely provides a snapshot of schools’ participation in the initiative. Future studies should employ multi-year data collection protocols to examine participation patterns and investigate factors that may be associated with variation in implementation rates and strategies. Furthermore, studies adopting experimental designs will be useful in determining cause-effect relationships between implementation practices and outcomes.

## Conclusions

This study provides an initial glimpse into schools’ implementation of Dubai 30×30 from a whole-of-school PA perspective. In line with recommended practices (CDC, 2019; IOM, 2013; Milton et al., 2021), physical education served as a central component in the promotion of PA within the participating schools and physical education teachers played leading roles in organizing and implementing Dubai 30×30 activities. While implementation resulted in several positive outcomes, physical education teachers bore the brunt of responsibility for schools’ participation in the initiative and a gap in PA promotion was evident during regular classroom time. Further research is needed to determine whether these results generalize to other schools in Dubai, and if so, how to optimally support collaborations between physical education teachers and other school stakeholders to harness Dubai 30×30 for whole-of-school PA.

## Data Availability

The data for this study are confidential as they focus on children and school professionals who participated in surveys, observations, interviews and focus groups.

## Acknowledgements

The authors wish to acknowledge the support of the principals, teachers and students who agreed to participate in this study.

## Author Contributions

CM and CAW conceived and designed the study. CAW, CEH, GT, ZMI, SZFN, MG, SK, MK, TK, PP and YS collected data for the study. CM and CAW organized the database. RGW performed the quantitative data analysis. CM and CAW performed the qualitative analysis. CM, CAW and RGW wrote the first draft of the manuscript. All authors contributed to manuscript revision, read, and approved the submitted version.

## References

Alrahma, A. M., Al Suwaidi, H., AlGurg, R., Farah, Z., Khansaheb, H., Ajja, R., … Loney, T. (2023). Results from the United Arab Emirates 2022 report card on physical activity for children and adolescents. Journal of Exercise Science and Fitness, 21(2), 218–225. 10.1016/j.jesf.2023.02.002

American Alliance for Health, Physical Education, Recreation and Dance. (2011). Comprehensive School Physical Activity Program (CSPAP) survey report. Reston, VA: Author.

Beighle, A., Erwin, H., Castelli, D., & Ernst, M. (2009). Preparing physical educators for the role of physical activity director. Journal of Physical Education, Recreation and Dance, 80(4), 24–29. 10.1080/07303084.2009.10598307

Belton, S., Britton, Ú., Murtagh, E., Meegan, S., Duff, C., & McGann, J. (2020). Ten years of “Flying the Flag”: An overview and retrospective consideration of the Active School Flag physical activity initiative for children – design, development and evaluation. Children, 7(12), 300. 10.3390/children7120300

Brener, N. D., Demissie, Z., McManus, T., Shanklin, S. L., Queen, B. and Kann, L. (2017). School Health Profiles 2016: Characteristics of health programs among secondary schools. Atlanta, GA: Centers for Disease Control and Prevention.

Bronfenbrenner, U. (1992). Ecological systems theory. In R. Vasta (Ed.), Six Theories of Child Development: Revised formulations and current issues (pp. 187–249). London: Jessica Kingsley Publishers.

Carson, R. L., Castelli, D. M., Beighle, A., & Erwin, H. (2014). School-based physical activity promotion: A conceptual framework for research and practice. Childhood Obesity, 10(2), 100–106. 10.1089/chi.2013.0134

Cassar, S., Salmon, J., Timperio, A., Koch, S., & Koorts, H. (2020). A qualitative study of school leader experiences adopting and implementing a whole of school physical activity and sedentary behavior programme: Transform Us! Health Education, 122(3), 267–285. 10.1108/HE-05-2020-0031

Centers for Disease Control and Prevention. (2013). Comprehensive school physical activity programs: A guide for schools. Atlanta, GA: U.S. Department of Health and Human Services.

Centers for Disease Control and Prevention. (2019). Increasing physical education and physical activity: A framework for schools. Atlanta, GA: U.S. Department of Health and Human Services.

Chaput, J. P., Willumsen, J., Bull, F., Chou, R., Ekelund, U., Firth, J., Jago, R., Ortega, F. B., & Katzmarzyk, P. T. (2020). 2020 WHO guidelines on physical activity and sedentary behaviour for children and adolescents aged 5-17 years: summary of the evidence. The International Journal of Behavioral Nutrition and Physical Activity, 17(1), 141. 10.1186/s12966-020-01037-z

Colabianchi, N., Griffin, J. L., Slater, S. J., O’Malley, P. M., & Johnston, L. D. (2015). The Whole-of-school approach to physical activity: Findings from a national sample of U.S. secondary students. American Journal of Preventive Medicine, 49(3), 387–394. 10.1016/j.amepre.2015.02.012

Edmonds, A. W., & Kennedy, T. D. (2017). An applied guide to research designs: Quantitative, qualitative and mixed methods. Los Angeles: SAGE Publications.

Global Advocacy for Physical Activity. (2011). Non-communicable disease prevention – investments that work for physical activity. International Society for Physical Activity and Health. Available from http://www.globalpa.org.uk/pdf/investments-work.pdf

Guthold, R., Stevens, G. A., Riley, L. M., & Bull, F. C. (2020). Global trends in insufficient physical activity among adolescents: A pooled analysis of 298 population-based surveys with 1·6 million participants. The Lancet. Child & Adolescent Health, 4(1), 23–35. 10.1016/S2352-4642(19)30323-2

Hayes, G., Dowd, K. P., MacDonncha, C., & Donnelly, A. E. (2019). Tracking of physical activity and sedentary behavior from adolescence to young adulthood: A systematic literature review. The Journal of Adolescent Health, 65(4), 446–454. 10.1016/j.jadohealth.2019.03.013

Schools get moving in the Dubai Fitness Challenge. (2022, December 1). Gulf Youth Sport. https://gulfyouthsport.com/2022/12/01/schools-get-moving-in-the-dubai-fitness-challenge/

Roberts, S. (2022, November 18). UAE schools embrace Dubai Fitness Challenge. Which School Advisor. https://whichschooladvisor.com/uae/school-news/uae-schools-embrace-dubai-fitness-challenge-30x30

Dubai Fitness Challenge: 88% participants hit 30-minute daily target last year. (2022, March 13). Khaleej Times. https://www.khaleejtimes.com/life-and-living/dubai-fitness-challenge-88-participants-hit-30-minute-daily-target-last-year#:~:text=The%20DFC%20has%20grown%20phenomenally,people%20take%20part%20in%202021

Katewongsa, P., Choolers, P., Saonuam, P., Widyastari, D. A. (2022). Effectiveness of a whole-of-school approach in promoting physical activity for children: Evidence from cohort study in primary schools in Thailand. Journal of Teaching in Physical Education, 4(2), 225–234. 10.1123/jtpe.2021-0186

Krueger, R. A., & Casey, M.A. (2014). Focus groups: A practical guide for applied research. New Delhi: Sage Publications.

Lincoln, Y., & Guba, E. G. (1985). Naturalistic inquiry. Newbury Park, CA: Sage.

Lonsdale, C., Sabiston, C. M., Raedeke, T. D., Ha, A. S., & Sum, R. K. (2009). Self- determined motivation and students’ physical activity during structured physical education lessons and free choice periods. Preventive Medicine, 48(1), 69–73. 10.1016/j.ypmed.2008.09.013

McIver, K. L., Brown, W. H., Pfeiffer, K. A., Dowda, M., & Pate, R. R. (2016). Development and testing of the Observational System for Recording Physical Activity in Children: Elementary School. Research Quarterly for Exercise and Sport, 87(1), 101–109. 10.1080/02701367.2015.1125994

McKenzie, T. L., & Lounsbery, M. A. (2013). Physical education teacher effectiveness in a public health context. Research Quarterly for Exercise and Sport, 84(4), 419–430. 10.1080/02701367.2013.844025

McLeroy, K. R., Bibeau, D., Steckler, A., & Glanz, K. (1988). An ecological perspective on health promotion programs. Health Education Quarterly, 15(4), 351–377. 10.1177/109019818801500401

Milton, K., Cavill, N., Chalkley, A., Foster, C., Gomersall, S., Hagstromer, M., … Schipperijn, J. (2021). Eight investments that work for physical activity. Journal of Physical Activity & Health, 18(6), 625–630. 10.1123/jpah.2021-0112

McMullen, J. M., Kallio, J., & Tammelin, T. H. (2022) Physical activity opportunities for secondary school students: International best practices for whole-of-school physical activity programs. European Physical Education Review, 28(4), 890–905. 10.1177/1356336X221092281

Moon, J., & Webster, C. A. (2019). MI (my) wheelhouse: A movement integration progression framework for elementary classroom teachers. Journal of Physical Education, Recreation and Dance, 90(7), 38–45. 10.1080/07303084.2019.1644258

Ní Chróinín, D., & McMullen, J. (2019) “The world is a happier place”: Celebration in a whole-of-school physical activity initiative. European Physical Education Review, 26(2), 337–352. 10.1177/1356336X19858115

Norris, E., van Steen, T., Direito, A., & Stamatakis, E. (2020). Physically active lessons in schools and their impact on physical activity, educational, health and cognition outcomes: a systematic review and meta-analysis. British Journal of Sports Medicine, 54(14), 826–838. 10.1136/bjsports-2018-100502

Orendorff, K., Webster, C. A., Mîndrila, D., Cunningham, K. M., Doutis, P., Dauenhauer, B., & Stodden, D. F. (2021). Principals’ involvement in comprehensive school physical activity programs: A social-ecological perspective. European Physical Education Review, 27(3), 574–594. 10.1177/1356336X20976687

Phelps, A., Calvert, H. G., Hwang, J., Glowacki, E., Carson, R. L., & Castelli, D. (2018). Environmental characteristics related to comprehensive school physical activity program implementation. European Journal of Environment and Public Health, 2(2), 07. 10.20897/ejeph/92007

Pirrie, M., Carson, V., Dubin, J. A., & Leatherdale, S. T. (2022). A comprehensive school health approach to student physical activity: A multilevel analysis examining the association between school-level factors and student physical activity behaviors. The Journal of School Health, 92(8), 774–785. 10.1111/josh.13178

Pulling Kuhn, A., Stoepker, P., Dauenhauer, B., & Carson, R. L. (2021). A systematic review of multi-component comprehensive school physical activity program (CSPAP) interventions. American Journal of Health Promotion, 35(8), 1129–1149. 10.1177/08901171211013281

Russ, L. B., Webster, C. A., Beets, M. W., & Phillips, D. S. (2015). Systematic review and meta-analysis of multi-component interventions through schools to increase physical activity. Journal of Physical Activity & Health, 12(10), 1436–1446. 10.1123/jpah.2014-0244

Schmidt, S. K., Bratland-Sanda, S. & Bongaardt, R. (2022). Secondary school teachers’ experiences with classroom-based physically active learning: “I’m excited, but it’s really hard”. Teaching and Teacher Education, 116, 103753. 10.1016/j.tate.2022.103753

Stoepker, P., Dauenhauer, B., Carson, R. L., & Moore, J. B. (2021). Comprehensive School Physical Activity Program Policies and Practices Questionnaire (CSPAP-Q). Research Quarterly for Exercise and Sport, 92(1), 100–110. 10.1080/02701367.2019.1711008

Strauss, A., & Corbin, J. M. (1990). Basics of qualitative research: Grounded theory procedures and techniques. Thousand Oaks, CA: Sage Publications.

Telama R. (2009). Tracking of physical activity from childhood to adulthood: A review. Obesity Facts, 2(3), 187–195. 10.1159/000222244

van der Mars, H. (1989) Observer reliability: Issues and procedures. In P. Darst,. D. Zakrajsek. and V. Mancini (Eds.), Analyzing Physical Education and Sport Instruction (2nd ed., pp. 53–80). Human Kinetics, Champaign.

Webster, C. A., Beets, M., Weaver, R. G., Vazou, S., & Russ, L. (2015). Rethinking recommendations for implementing comprehensive school physical activity programs: A partnership model. Quest, 67, 185–202. 10.1080/00336297.2015.1017588

Webster, C.A., Caputi, P., Perreault, M., Doan, R., Doutis, P. and Weaver, R.G. (2013) ‘Elementary classroom teachers’ adoption of physical activity promotion in the context of a statewide policy: An innovation diffusion and socio-ecologic perspective. Journal of Teaching in Physical Education, 32(4), 419–440. 10.1123/jtpe.32.4.419

Webster, C. A., Rink, J. E., Carson, R. L., Moon, J., & Gaudreault, K. L. (2020). The comprehensive school physical activity program model: A proposed illustrative supplement to help move the needle on youth physical activity. Kinesiology Review, 9(2), 112–121. 10.1123/kr.2019-0048

World Health Organization. (2022). Promoting physical activity through schools: Policy brief. Available from https://apps.who.int/iris/bitstream/handle/10665/354605/9789240049567-eng.pdf?sequence=1

